# Use of Systems Thinking and Group Model Building Methods to Understand Patterns of Continuous Glucose Monitoring Use Among Older Adults with Type 1 Diabetes

**DOI:** 10.1101/2022.08.04.22278427

**Authors:** Anna R. Kahkoska, Cambray Smith, Laura A. Young, Kristen Hassmiller Lich

## Abstract

A growing number of older adults (ages 65+ years) live with Type 1 diabetes, yet little is known about the complex dynamics that promote use of diabetes technology, such as continuous glucose monitoring (CGM), in this age group. We used systems thinking and methods from group model building (GMB), a participatory approach to system dynamics modeling, to collect data from older adults with Type 1 diabetes and their caregivers through group workshops and individual validation interviews. Data were integrated into a causal loop diagram of the “system” of factors associated with CGM uptake and use, including the clinical and psychosocial outcomes of use and interactions with caregiver and healthcare system factors. We describe the study design, recruitment, GMB and interview procedures, participant feedback, and lessons learned. The study demonstrates feasibility, acceptability, and the value of GMB to engage older adult stakeholders in sophisticated and rigorous research about key determinants of complex health outcomes over time.

## Introduction

Type 1 diabetes is a chronic disease in which the pancreas no longer produces insulin, the hormone critical for blood glucose homeostasis (1). Exposure to elevated blood glucose levels (i.e., hyperglycemia) over time is associated with the development multiple chronic complications, including neuropathy, retinopathy, nephropathy, and cardiovascular disease, while episodes of low blood sugar (i.e., hypoglycemia) can be life-threatening and require urgent attention (1, 2). As a result, constant self-management is required to maintain blood glucose levels as near-normal as possible. Unfortunately, self-management is made challenging by dynamic insulin needs, which can be influenced by dietary intake, physical activity, stress, and illness, and thus vary hour-to-hour, day-to-day, and over time (2). As a result, individuals living with Type 1 diabetes are tasked with measuring their blood glucose levels, assessing and accounting for dietary intake, dosing and timing exogenous insulin delivered through injection or insulin pump modalities, responding to hyper- and hypoglycemia, and accounting for other factors such as physical activity, stress, and illness (2).

### Older Adults with Type 1 Diabetes

As the US older adult population (≥65 years) grows and the life expectancy associated with a diagnosis of Type 1 diabetes increases, a sizable population of older adults living with Type 1 diabetes has emerged; this population is expected to continue expanding in upcoming years (3). From a clinical perspective, care and management of Type 1 diabetes in older adulthood is often complex, as patients vary according to age, functional health, presence of frailty, and comorbidity profiles (4). Compared to younger adults living with Type 1 diabetes, for whom the primary focus of care and self-management is on robust glucose control, older adults living with Type 1 diabetes should primarily be focusing on the avoidance of hypoglycemia. Older adults have an increased risk for hypoglycemia, which remains a grave clinical concern due to high morbidity and mortality (4-6). Although ensuring patient safety through the avoidance of hypoglycemia, accommodating patient preferences, and preserving quality of life have been outlined as objectives for care (7-10), more specific and applied data to guide care in this population are currently scant, largely owing to relatively recent emergence of this patient population (4, 5, 10).

### Technologic Approaches for Type 1 Diabetes Management

New technologic approaches for both glucose monitoring and insulin delivery have been developed to improve strategies for Type 1 diabetes management (2). One such development is continuous glucose monitoring (CGM), a remote monitoring approach to blood glucose measurement. CGM systems include three components: an on-body sensor with a subcutaneous catheter to measure interstitial glucose approximately every five minutes, a Bluetooth “transmitter,” and an external receiver which displays the real-time blood glucose (11, 12). CGM is offered currently in two forms, including real-time CGM, or systems that measure and display real-time or near real-time glucose levels at all time, and intermittently scanned CGM, systems that require individuals to scan their device against the sensor to access glucose information (12). Both types of CGM, offer several major advantages over alternative, invasive self-monitoring approaches for blood glucose, which require individuals to obtain a small blood sample *via* finger prick and use a glucometer to measure glucose levels therein. These advantages include access to real-time or near-real time blood glucose information, data on glucose trends (including the rate of rising and falling glucose levels), and less invasive testing methods. Based on growing evidence to suggest clinical and patient-oriented benefits of CGM use, including improved glycemic control and psychosocial wellbeing, clinical practice guidelines now suggest that CGM be offered for all adults with Type 1 diabetes (12). Practice guidelines specify that adults with diabetes must be capable of using CGM themselves, which may include help from a caregiver, and the specific selection of device should reflect individual patient circumstances, preferences, and clinical needs (12).

### Benefits and Challenges of Continuous Glucose Monitoring for Older Adults with Type 1 Diabetes

Despite the advantages, a major knowledge gap specifically exists regarding how older adults with Type 1 diabetes interact with, and may ultimately benefit from, diabetes technology like CGM. This gap was highlighted as a critical area for future research in a 2020 consensus statement published on behalf of the International Geriatric Diabetes Society (4). Data from efficacy-based studies suggest that CGM may confer a significant safety benefit for this age group; in a randomized control trial, use of CGM reduced hypoglycemia over six months among older adults with T1D (13). The trial measured the duration of hypoglycemia, or the time that blood glucose levels were below 70 mg/dL, at baseline and over six months of CGM use (13). The group who was randomized to use CGM had a reduction in hypoglycemia from approximately seventy-three minutes per day at the trial baseline to thirty-nine minutes per day over the six-month trial period; there were no significant changes in the duration of hypoglycemia among the group who was randomized to receive standard of care only (13). Importantly, the reduction in hypoglycemia occurred concurrently with improvements in overall glycemic control, as measured by hemoglobin A1c as well as the time-in-range, or duration of time that blood glucose levels were measured between 70 mg/dL and 180 mg/dL (13). This finding was important in showing that the reduction in hypoglycemia did not come at the cost of more time spent in hyperglycemic ranges. A handful of observational studied have further reinforced positive effects of CGM in older adults, including decreased hypoglycemia (14), reduced hemoglobin A1c and glycemic variability (15), and increased well-being and feelings of security (16).

Although estimates of the prevalence of CGM use in real-world populations of older adults vary, they range between approximately 30-70% in various studies and thus suggest opportunities to increase uptake (14, 17, 18). It is further established that the general use of medical technology may represent a complicated issue for older adults, particularly with regards to unique, age-specific barriers and the range of biopsychosocial needs that exist across the population (19). For example, physical symptoms, functional limitations, barriers to care, and psychosocial wellbeing all impact on disease self-management and may impact technology uptake. The growing number of chronic medical conditions accrued in older adulthood lends further complexity to integrating tools that may help improve quality of life. Accessibility features are lacking, including those to address changes to dexterity, visual acuity, and hearing loss (19). From a psychosocial perspective, older adults may find learning new technologies to be challenging (14) and may require more time for education and training to use CGM and learn to interpret data (17). Compared to younger adults, studies using questionnaire data have shown older adults perceive substantially higher burdens of technology such as CGM, including concerns that sensor readings cannot be trusted, information from CGM may cause too much worry, and that the technology will be too hard to understand (17). Interestingly, differences in perceived burdens were substantially less pronounced across age groups in those who use CGM, suggesting that with adequate time, training, and support, older adults can use CGM effectively and experience clinical benefits (13, 15, 17). ***However, complex interventions such as this are often plagued with challenges (20), and so identifying the most critical elements to support success as efficiently as possible will increase the likelihood of successful translation across the broader population***.

### Objective of the Study and Selection of the Research Approach

We focused on the following gap in the literature: The experiences of older adults with Type 1 diabetes with CGM have not been well-characterized (4). For example, there are very limited data on what supports uptake of CGM from the perspective of patients and their caregivers, how this technology impacts disease self-management, lived experiences, and clinical outcomes, and what suboptimal responses to technologic approaches over time may occur and why.(4, 21) The objective of this study was to understand the complex nature of older adults’ experiences associated with initiating and sustaining use of CGM, including changes in different clinical, behavioral, and psychosocial variables over time, and how these variables interact to ultimately produce patters of effective use versus less effective use or nonuse. These data are needed to inform how clinical recommendations and supports can be developed to help all individuals with Type 1 diabetes incorporate the ever-evolving technologic aspects of diabetes management into their care regimes, regardless of biologic, clinical, and psychosocial differences (22). As the population of older adults with diabetes grows, these data are also needed to ensure existing and emerging diabetes technology remain accessible across the lifespan.

From a scientific perspective, a major challenge is how to capture the multiple, interrelated factors that may shape older adults’ experiences with diabetes self-management and technology use, as well as the dynamic complexity of those factors. Dynamic complexity refers to situations in which effects of time are not easily explained through simple cause and effect, but rather represent the influences from multiple interacting factors that may be non-linear, occur over variable durations of times, and trigger powerful feedback loops to reinforce or balance changes within the web of interconnections (23-25). While survey items may limit the breadth of response options possible, even traditional qualitative data collection, such as focus group or key informant interviews, may not offer insight into complex patters of change, sequenced events, or feedback loops between behaviors and outcomes. Instead, we hypothesized that a systems thinking approach is well-suited for this task, offering methods that can capture multiple perspectives and elucidate complex dynamics of systems (here, those affecting CGM use) in a more holistic way (26, 27). A system is the set of interconnected elements that interact with each other to produce emergent effects, or outcomes we care about (i.e., want to reinforce or change) (28). These effects persist over time and adapt to changing circumstances (26). In the context of lived patient experiences, such as those had by older adults as they learn to use CGM for their diabetes management, that system could include factors such as physical symptoms or clinical outcomes, lifestyle and behavioral aspects of disease management, wellbeing and psychosocial changes, as well as individual preferences, social and environmental forces, and healthcare resources. We thus explored how participatory system dynamics methods could be leveraged to elucidate and document the multiple, interconnected variables that interrelate to produce several emergent and distinct patient profiles of experience with CGM use over time (26, 27).

We applied group model building methods to collect data from older adults living with Type 1 diabetes and their caregivers, with considerations to accommodate logistical constraints (e.g., participants bringing heterogeneous clinical, personal, and professional backgrounds, limiting the study duration to no more than three hours participants initially). Group model building is a participatory approach to system dynamics in which diverse stakeholders can exchange their perceptions and experiences to collectively consider the causes of a dynamically complex problem (25, 29-31). With this approach, we aimed to bring a systems thinking framework and system dynamics techniques to represent and model the complex processes and outcomes of older adults initiating and using CGM over time and to uncover factors relating to sustained and effective use in daily life. To our knowledge, no group model building studies have yet been published that adapt these methods to specifically involve older adults in improving clinical care by developing a better understanding of broader forces affecting clinical outcomes; this a thus rich perspective otherwise missing from geriatric research. As part of this study, we therefore also explored how systems thinking could be taught to older adult research participants and group model building methodology could be leveraged to engage stakeholders to identify key pathways where effective therapy use and self-management break down, elucidate the problematic outcome trajectories, reconcile incentives and constraints of real-life care and support systems, and identify opportunities for change that are aligned with their experienced system structure.

## Materials and Methods

### Study Design

We developed a facilitation guide and applied it within a series of small, parallel group model building workshops to understand older adults with Type 1 diabetes’ experiences initiating and using CGM over time. The study included two main components: a series of three-hour, in-person, small-group group model building workshops and a follow-up series of one-on-one virtual validation interviews. Upon completion of the study, all participants received a $100 (USD) incentive for their time and effort. All study procedures were approved by the Institutional Review Board at the University of North Carolina at Chapel Hill.

### Study Participants

#### Eligibility Criteria

For the in-person workshops, patient participants were eligible if: they had a diagnosis of Type 1 diabetes documented in the electronic medical record, were ≥65 years of age at the time of recruitment, used an insulin regimen of pump or multiple daily injections, were able to manage their diabetes independently or with the help of a caregiver, had a Hemoglobin A1c level measured within the past year ≤10.0%, and comprehended written and spoken English. Patients were eligible to participate regardless of CGM use status or the brand of CGM device used. Participants were ineligible if they had a significant medical or psychiatric condition that may prohibit completion of the workshop, a clinical diagnosis of dementia, or were not fully vaccinated against COVID-19 at the time of recruitment (increasing risk of transmission in in-person sessions). All potential patient participants were invited to bring a caregiver with them to the research study. Caregiver participants were eligible for the study if they were invited by participants living with diabetes and serve a ‘caregiver’ role in the sense that they provide daily or regular care or support with regards to specific aspects of care or daily management for an older adult (≥65 years) with Type 1 diabetes.

#### Recruitment

Patients were recruited from the outpatient diabetes clinic at an academic medical center. For the in-person workshops, potentially eligible participants were identified via the electronic health record system and contacted *via* email and telephone outreach. All interested participants were ultimately contacted by telephone following a standardized recruitment script in which participants were provided information about the study and invited to optionally bring a caregiver to the workshop. CGM use status and vaccination status were determined by chart review and confirmed verbally as part of recruitment. Participants were scheduled for a workshop on a rolling basis and provided with a series of confirmation and reminder emails.

All participants of the in-person workshop were invited to participate in validation interviews; they indicated their preference in writing at the close of the workshop and provided an email address for further contact.

### Group Model Building Procedures

Each in-person workshop followed a uniform structure including: 1) completion of a brief questionnaire; 2) the group model building session, including both didactic components and structured activities; and 3) a brief focus group discussion. The workshop lasted three hours in duration. A sample agenda is shown in **Table 1**.

**Table 1.**
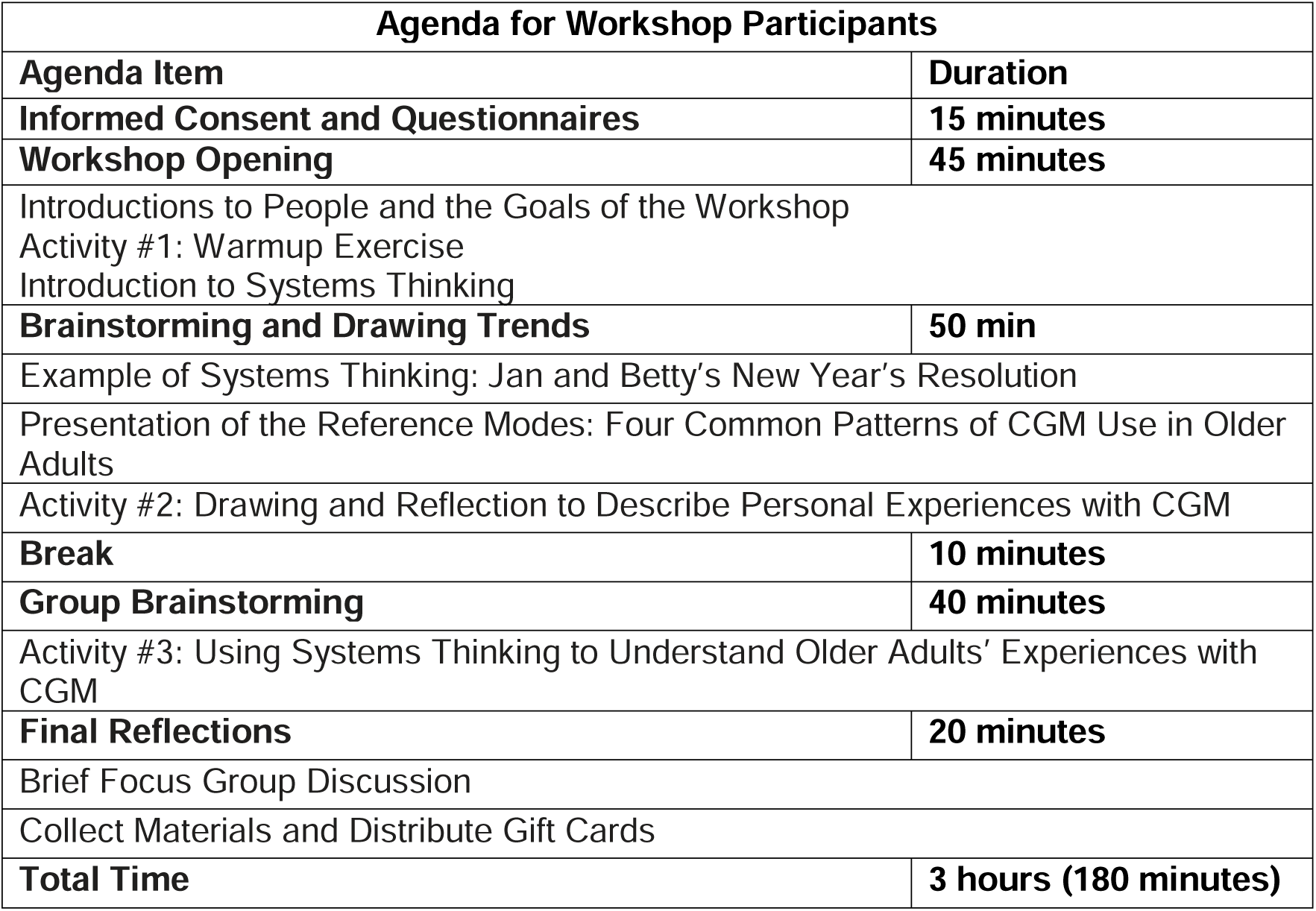
Sample group model building workshop agenda. The agenda included the following note for study participants: This is an interactive workshop where we will essentially brainstorm on the wide range of experiences that older adults may have, as well as the different underlying factors that might lead to those experiences. The agenda below is a guide, but this is a fluid workshop. We will take extra breaks as necessary.

| **Agenda for Workshop Participants** |  |
| --- | --- |
| **Agenda Item** | **Duration** |
| **Informed Consent and Questionnaires** | **15 minutes** |
| **Workshop Opening** | **45 minutes** |
| Introductions to People and the Goals of the Workshop<br>Activity #1: Warmup Exercise<br>Introduction to Systems Thinking |  |
| **Brainstorming and Drawing Trends** | **50 min** |
| Example of Systems Thinking: Jan and Betty's New Year's Resolution |  |
| Presentation of the Reference Modes: Four Common Patterns of CGM Use in Older Adults |  |
| Activity #2: Drawing and Reflection to Describe Personal Experiences with CGM |  |
| **Break** | **10 minutes** |
| **Group Brainstorming** | **40 minutes** |
| Activity #3: Using Systems Thinking to Understand Older Adults' Experiences with CGM |  |
| **Final Reflections** | **20 minutes** |
| Brief Focus Group Discussion |  |
| Collect Materials and Distribute Gift Cards |  |
| **Total Time** | **3 hours (180 minutes)** |

The research team included one facilitator and one research assistant. The workshop was held in a moderate sized-conference room at a clinical research building. The room included a large table, up to 12 chairs, a projector with HDMI connector cables, and a screen at the front of the room. To ensure safety for participants in the in-person workshop, all team members and participants were fully vaccinated as per exclusion criteria above, practiced adequate spacing (e.g., six feet between participants), and remained masked for the entirety of the session. Each study participant was provided with an assigned seat and provided with a clipboard that contained the consent form and HIPAA authorization, the questionnaire, and the workshop packet. Each participant seat at the table was marked, and there were multiple black and colored pens, two individual whiteboards and colored markers, and two small pads of sticky notes provided. Due to COVID-19 and the need to maintain masking, no beverages or food were provided, although participants were encouraged to take breaks to eat and drink as needed.

The workshop opening involved open-ended prompts for introductions, an icebreaker, and sufficient time for study participants to interact and build rapport before starting the structured aspects of the workshop, so that they would feel comfortable sharing their views and brainstorming in a group. Following brief introductions of the research team, participants were asked sequentially to introduce themselves and describe, to the extent they were comfortable, their relationship to Type 1 diabetes. The introduction prompt was selected to allow for a range of possible responses, which may include narratives surrounding diagnosis stories, experiences with changing treatment regimens or self-management, and attitudes towards Type 1 diabetes. A separate, informal icebreaker was chosen for each workshop, including, “What is your favorite Thanksgiving food?” and “What is your favorite ice cream flavor?” Although icebreakers are not a requisite aspect of group model building, providing participants with an open structure to tell their stories and allowing time for reactions from other participants early in the workshop aimed to facilitate group bonding to support the rest of the workshop activities.

#### Didactic Component

Each workshop opened with a short presentation by the facilitator that included an overview of the rationale for the study, the goals for data collection, and a series of “ground rules.” The ground rules encouraged participants to share their thoughts freely, to draw upon their experiences as the ‘experts’ in the room, and take breaks as needed. An iceberg metaphor was used to introduce the concept of systems thinking (32), in which isolated events were framed as the ‘tip’ of the iceberg, while related and concerning trends (e.g., root causes, reference modes that put the events in context over time) and problematic aspects of underlying system structure and mental models were reflected as the part of the iceberg that was below the waterline (**Figure 1A**). Of note, the “Iceberg Model” represents a ubiquitously used image to teach systems thinking by linking events to patterns to system structure (33).

**Figure 1.**
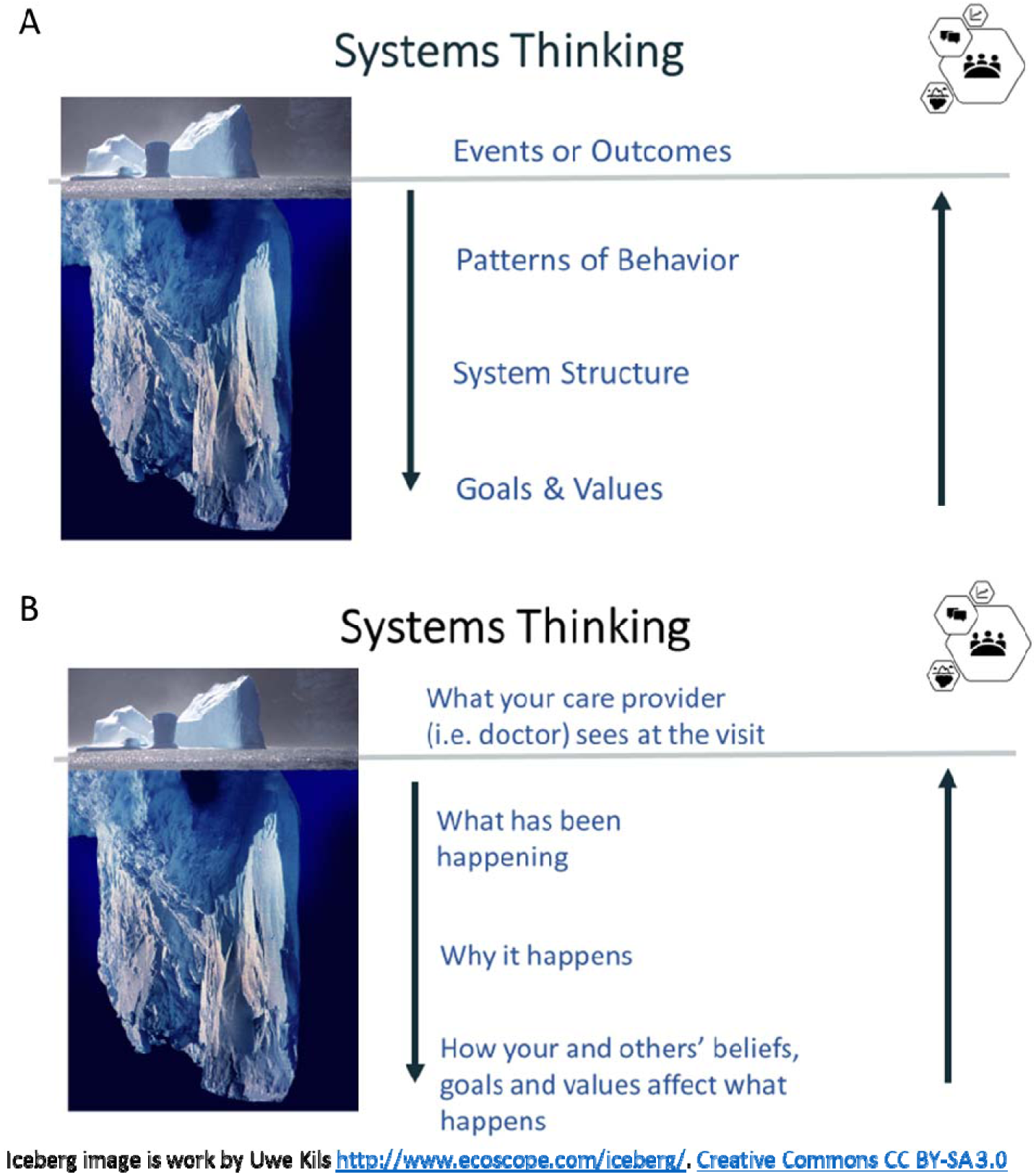
Didactic study components used to present systems thinking. Panel A shows the general framework, while Panel B shows its extension to understand Type 1 diabetes self-management experiences (B). Note: the ‘Iceberg Model’ (32) is a widely used approach for teaching introductory systems thinking systems thinking (33). The iceberg image is work by Uwe Kils. http://www.ecoscope.com/iceberg/. Creative Commons CC BY-SA 3.0

The iceberg metaphor was then extended from a general framework to apply to Type 1 diabetes self-management, in which participants were invited to help the research team understand the experiences that happen “below the waterline” as it relates to initiating and using CGM.

The facilitator provided an example of how the systems thinking framework would be applied, which focused on two hypothetical older adult characters in a related but not too overlapping example. In the example, the characters were friends who set the same New Year’s Resolution to walk 10,000 steps per day and had different outcomes over the following six months. The example was used to introduce the concept of reference modes, behavior-over-time graphs to describe the related trends, and causal loop diagramming. The example and didactic language are presented in full in **Appendix A: Primer to Systems Thinking and Systems Mapping**.

#### Reference Modes

Following the example of systems thinking, the rest of the workshop focused on CGM use in older adults. We selected four reference modes, or real-world patterns of behavior over time, to reflect common trajectories of optimal and suboptimal CGM use over the first six months following initiation of therapy (i.e., consistently high use, moderate use increasing to high use, continually declining use, and intermittent use/oscillation). We aimed to present enough different reference modes to capture the breadth of common real-world use patterns, while avoiding excessive or redundant trends which may contribute to participant fatigue (i.e., we strived to illustrate the smallest set of distinct reference mode shapes that would elucidate the breadth of qualitatively distinct feedback structures). Each reference mode was presented as a hypothetical older adult character, which were used to introduce a 6-month behavior-over-time graph of CGM (see **Figure 2**). We defined CGM use (i.e., Y-axis) as both wearing the CGM and using the readings to make decisions for Type 1 diabetes management, such as ingesting carbohydrates or dosing insulin. Throughout the study, the research team referred to the reference modes by name of the corresponding older adult character – with the goal of understanding each common behavior-over-time profile. For each reference mode, we strove to draw out stories about key feedback loops operating at different points of time as described in Figure 2 (e.g., the reinforcing loop that might drive use up or down; balancing loops that counteract change – either limiting improvement or counteracting undesired drops in use).

**Figure 2:**
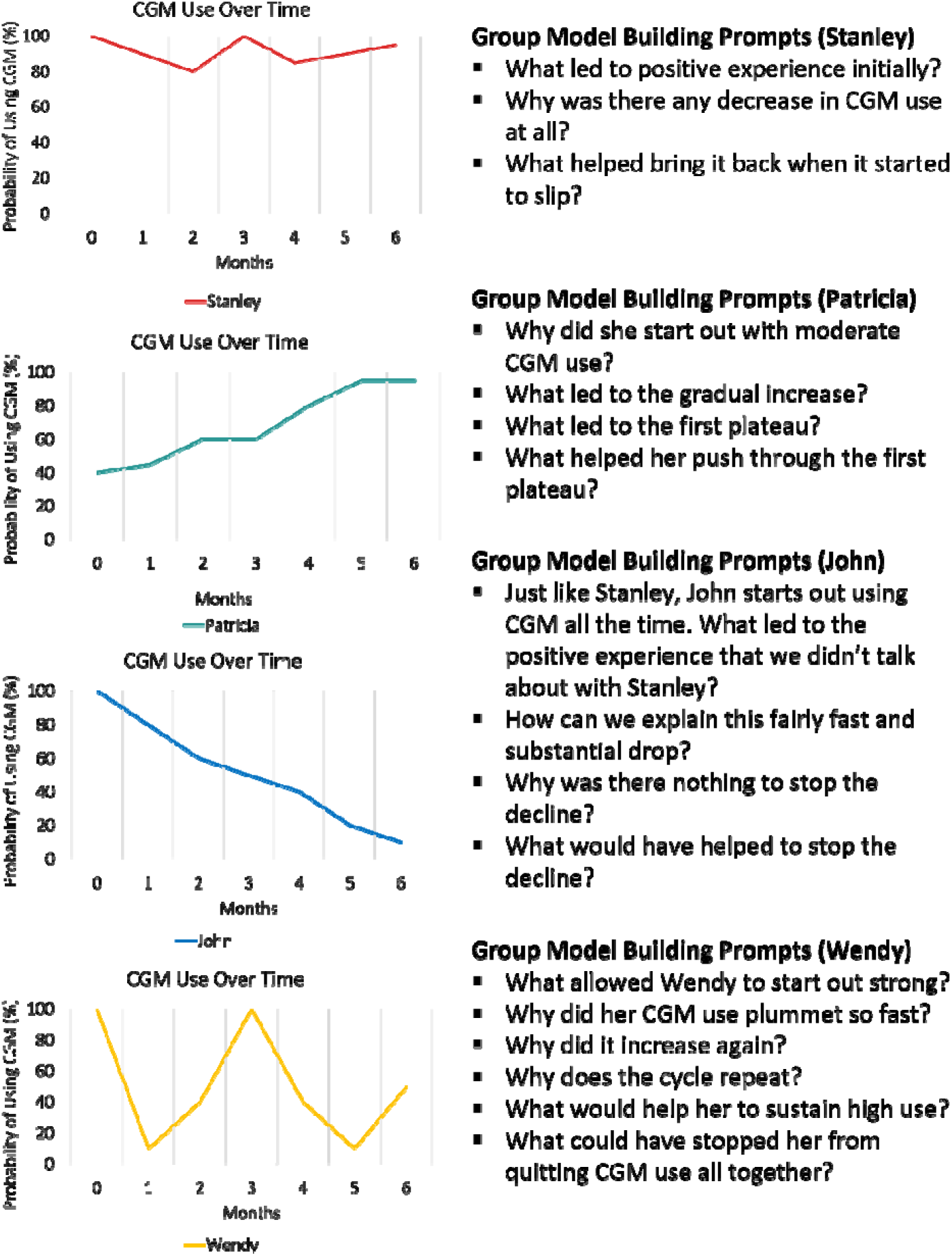
Reference modes provided during the group model building workshop. The reference modes were presented as named characters representing older adults living with Type 1 diabetes who began using CGM as part of their diabetes management. The graphs show the probability of CGM use, defined as both wearing the monitor and using glucose information for diabetes management, over the first six months of using CGM. Four reference modes were selected, including one to represent consistently high use (Stanley), moderate use increasing to high use (Patricia), continually declining use (John), and intermittent use (Wendy).

#### Behavior over time graphs

Following presentation of the reference modes, the workshop transitioned to drawing and group discussion activities. Behavior over time graphs were presented as graphs that focus on patterns of change over time, rather than on an isolated event or outcome, to help people and researchers think about how and why that change is happening. The facilitator introduced the concept of related trends, including guidelines for drawing and annotating trends, and suggested trend topics. Guidelines for brainstorming related trends included: 1) there are no right or wrong answers; 2) trends typically represented nouns or something that can increase or decrease over time unambiguously; 3) there is no need for a formal scale or measurement (i.e., it could be numbers/a specified range or qualitative – low to high); and 4) trends can be consequences or causes of CGM use over time. The facilitator presented an example of how to draw a graph over time, carefully labeling the X-axis as “Time,” noting the start time and end time. The Y-axis was labeled with a variable name and scale, and the understood trend(s) was (were) drawn on the graph and annotated (i.e., reasons the trend changed at specific points in time were noted).

Over the course of the pilot study, we experimented with a range of approaches to encourage the drawing of behavior over time graphs. In the first four workshops, study participants were invited to use their Workbook Packet or personal whiteboards to draw their own CGM use patterns and related trends. In the latter five workshops, participants were asked to use their Workshop Packet to identify which reference mode best reflected their own CGM use pattern. Former-users were asked to select the graph which represented their experience, while never-users were asked to select their imagined experience. Participants were then asked to identify and draw three emotions and three benefits that changed over their first six or more months of using CGM. A sample workshop packet from the latter five workshops can be found in **Appendix B: Group Model Building Workshop Packet**.

In any workshops, the facilitator answered questions, clarified tasks, and encouraged participants to ask for help if they experienced confusion. If participants were unable to draw themselves, members of the research team offered to listen to their stories and draw behavior over time graphs on their behalf. Following drawing exercises, the facilitator led a discussion in which each participant was asked to share their drawings through storytelling and to react to other participant’s drawings and stories.

#### Collective Annotation of the Reference Modes

To collect data for causal loop diagrams to model the system structure underlying common CGM use patterns, we applied a facilitated group model building process based on published scripted group exercises (30, 34).

For each reference mode, the facilitator posed a series of open-ended questions meant to uncover key associations and feedback loops to be represented within the causal loop diagram. Feedback loops are closed chains of causal connections, which can be reinforcing (i.e. when change in the first factor causes a series of changes that ultimately loop back to drive further change in that factor) or balancing (i.e. when change in the first factor causes a series of change that ultimately loop back to counteract the effects of the initial change) (28, 35-37). While reinforcing feedback loops can cause exponential growth or decline, balancing loops seek equilibrium within systems; feedback loops may have variable time delays (35, 37).The probing questions are shown in **Figure 2** with the corresponding reference modes. The reference modes were displayed on 24-inch x 36-inch laminated posters around the room, and participants’ ideas were scribed onto small sticky notes and used to annotate the diagram. The facilitator highlighted the feedback thinking for all four reference modes. At the point where a feedback chain became closed, the research team checked with the entire group to see if the chain was correct and complete. Throughout, participants were encouraged to brainstorm together and react to ideas across the group.

Throughout data collection, the research team periodically assessed saturation of themes proposed during the collection annotation of the reference mode. Saturation was defined as the point when no new or original themes emerged. Further recruitment was halted when saturation was achieved.

### Other Data Collection

#### Focus Group Discussion

Participants were engaged in a brief focus group discussion at the end of the workshop to provide a final opportunity for sharing thoughts about CGM use in older adults. The focus group discussion was guided by the following four questions: 1) We just talked through four examples here today. Can you think of a story of CGM use over time (i.e., a new reference mode) that we haven’t talked about? 2) With all of this in mind, what do you think are the top three things that we should know, study, or change to help older adults have positive experiences using CGM? 3) When you think about the things you do to take care of your diabetes every day, what are the ways that CGM can help the most? 4) What are expectations and goals that caregivers, doctors, and other members of the care team could have that would be supportive for older adults when they use CGM?

### Feedback on the Research Study

At the close of the workshop, participants were asked to use their Workshop Packets to provide feedback on the GMB session to understand how the group model building methodology was perceived among older adults with Type 1 diabetes. Participants were asked to rate their comfort level sharing all their experiences and thoughts (Likert scale; 1-5) and offered an opportunity to share in writing anything additional with the research team that they did not feel comfortable sharing with the group. Participants were also provided space to indicate what they liked and did not like about the workshop. Finally, they were provided with a brief ‘primer’ on systems thinking for optional take-home reading, which reinforced didactic comments from the workshop and included additional information about causal loop diagrams (see **Appendix A: Primer to Systems Thinking and Systems Mapping**.).

Workshop packets were collected and scanned. Photography was used to capture individual and group drawings that occurred outside of the packets, as well as the collective annotations of the reference modes. Workshops were audio-recorded and transcribed. All data were de-identified for analysis.

### Causal Loop Diagramming

Given overlap in variables generated through group model building across the four reference modes, the research team consolidated and merged data from each reference mode into one collective diagram depicting the factors, experiences, outcomes, and events that may interact to drive optimal versus suboptimal CGM use patterns over time. As the goal of understanding lived experiences relating to CGM use, we established a system boundary as factors intrinsic to a patient, in a patients’ life (home, social, etc.), or their clinical care environments shaping their CGM use. We designed a core structure to capture factors relating to uptake of CGM and ongoing use of CGM, as well as a subset of ‘dynamic’ drivers of CGM use, or factors those participants described as reinforcing or inhibiting CGM use over time that were directly modulated by experiences with the technology. Given that the same causal loops emerge differently in different narratives (depending on if they are driving CGM use or not), we focused on identifying the directionality of each causal association within a loop and the impact of the loop on dynamic CGM use. In cases where participants’ direct language was deemed to be the most accurate representation of a sentiment or concept in the map, *in vivo* factors were used.

### Validation of the Diagram

A key component of stakeholder-engaged systems sciences involves iterative refinement and updating of system models with new or changing information.(38) As explicit diagramming was not a part of the in-person workshop, we elected to validate our diagram in follow-up interviews. Participants of the in-person workshop were offered the opportunity to review final causal loop diagram components and offer their feedback through an individual, virtual follow-up interview. The objective of this validation scheme was to ensure that diagrams retained fidelity to the raw data and lived experiences of study participants. Because the full causal loop diagram included many variables and feedback loops, one component of the map was validated in detail with different participants. Validation interviews were 30-minutes and followed a standardized script including a brief overview of the objectives of the research study, a narrative study of main findings, a viewing of the full causal loop diagram, and an explanation of a focused, simplified view of the diagram or one subset of the model on which the validation interview focused. Feedback was structured around the following questions: 1) *What are your reactions to the full system map?* 2) *What part of [focused diagram] resonates most?* 3) *What pieces of [the focused diagram] are the most important?* 4) *What pieces of [the focused diagram are missing? This may include making changes such as adding factors, removing factors, or drawing new connections between factors*. Participant feedback was scribed. Validation interviews were performed as dyadic interviews when caregivers are present. The CLD was revised iteratively over the course of conducting interviews.

## Results

We adapted group model building methods, a participatory approach to system dynamics, to model experiences and trajectories of CGM use among older adults with Type 1 diabetes. We completed nine, in-person group model building workshops with older adult patients and their caregivers and generated an integrated, complete causal loop diagram. An in-depth description of study participants and the resulting causal loop diagram are provided elsewhere [manuscript *in prep*.] Herein, to characterize this novel approach to data collection in older adults, we illustrate data collected through each component of the GMB process, as well as other evaluation measures including recruitment outcomes and feedback on the study from participants.

### Recruitment and Attendance

Study recruitment spanned from November 10, 2022 – December 13, 2022. Nine workshops were held between November 15, 2022, and December 15, 2022. Each workshop had between two and six participants. A total of 33 older adults and caregivers participated, of which four were caregivers and the rest were individuals living with Type 1 diabetes. The mean age of the sample was 73.3±4.3 year, with a range of 66-85 years. 55% identified as women, 82% identified as non-Hispanic white, 12% were non-CGM users.

During recruitment, the main reasons cited for lack of interest or inability to participate included competing medical or surgical appointments, conflicts relating to the winter holidays, and non-local temporary or permanent residence. There were two major challenges for recruitment of an adequately diverse sample. First, the majority of eligible participants for the study within the medical center were non-Hispanic White race and ethnicity, and there was comparatively a very limited pool of Black or African American, Asian, American Indian or Alaska Native, native Hawaiian or Other Pacific Islander, or Hispanic participants for potential recruitment. The imbalanced recruitment pool was reflected in a study sample that was majority White race and non-Hispanic ethnicity. Second, there was a small number of eligible participants who were not CGM users, resulting in the majority of study participants being active CGM users. Attendance of the workshop by recruited individuals was relatively high. A total of four participants did not show for their scheduled workshop. One of those participants was rescheduled for a subsequent workshop, two were unable to be rescheduled and one was not successfully re-contacted.

### Behavior over time graphs

Individual drawing activities proved to be the most challenging aspect of the workshop for study participants, particularly when the drawing prompts were left open-ended. Approaches that facilitated older adults drawing included providing an example of a drawing, pairing a participant with a research team member to diagram on their behalf and in response to their storytelling, and providing more specific prompts, such as asking for graphs of named emotions associated with using CGM or benefits yielded over the first six month. **Figure 3A** depicts a sample of drawings of emotions and benefits from study participants, including both users and non-users of CGM. When not all study participants felt comfortable drawing, we found that those who did tended to lead storytelling, which revealed complex dynamics, catalyzing rich group discussions that were captured in the study transcripts and coded for inclusion in the causal loop diagram. These discussions also contributed to a significant amount of group bonding, particularly when participants found resonance in their drawings or stories.

**Figure 3.**
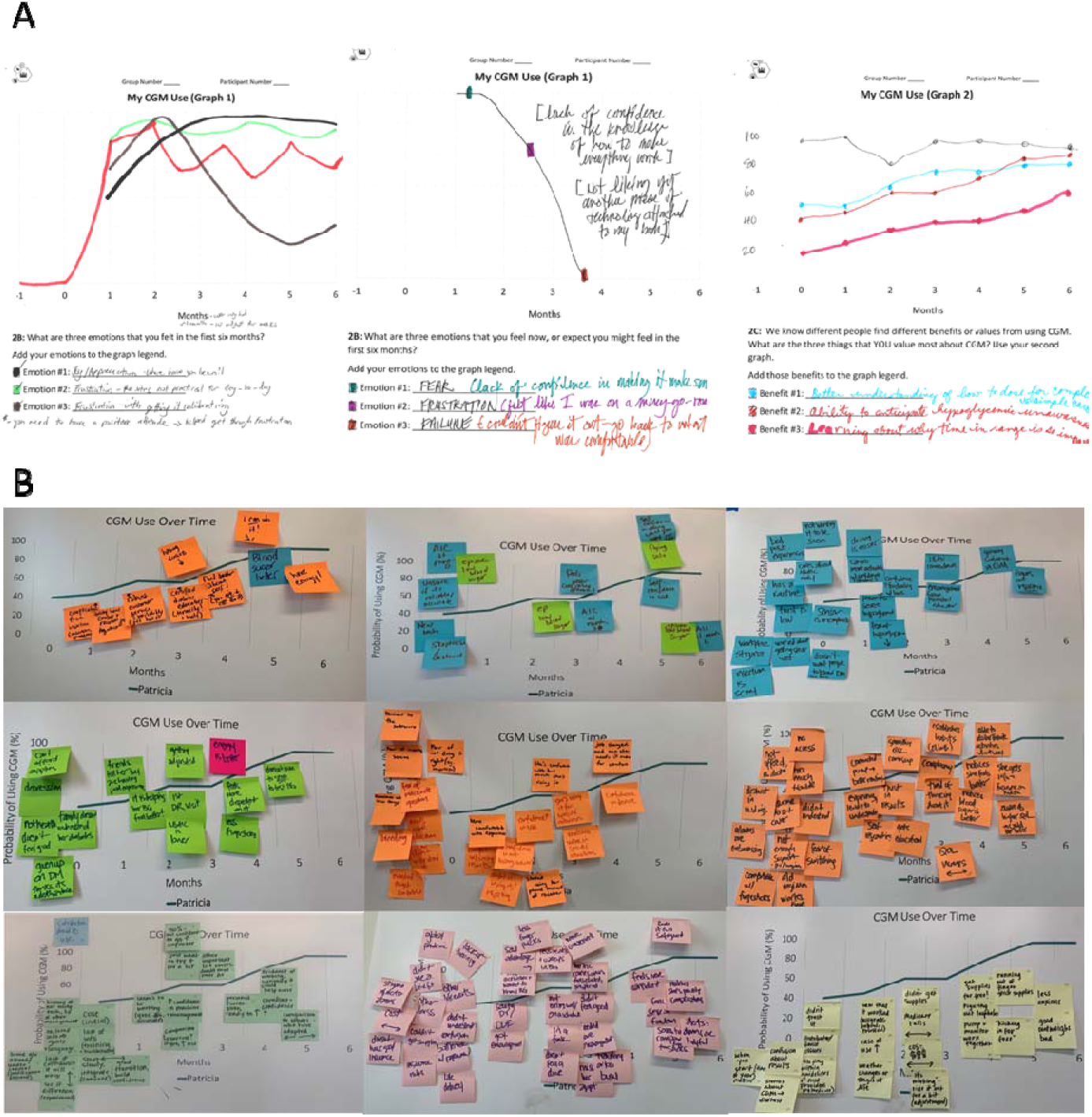
Selected illustrative examples of raw data generated as part of the in-person group model building workshop. Panel A shows a subset of individual behavior over time graphs drawn by older adults living with Type 1 diabetes and their caregivers. Panel B shows the collective annotations of the moderate use increasing to high use reference mode.

### Collective Annotation of the Reference Modes

Workshop participants were successfully engaged through a series of discussion questions to elicit information necessary for causal loop diagramming. **Figure 3B** shows an example of the raw data produced by collective annotation of the reference modes. Although some themes were constant across groups, different groups generated unique collective ideas, resulting in rich heterogeneity in the annotations across groups. Saturation was achieved in the themes of group annotations by the eighth workshop.

### Study Feedback

Participants expressed that they felt comfortable sharing their thoughts and experiences in the group format, where the mean comfort level was rated as 4.97 on a scale of 1-5 (5 is the highest; n=26 respondents), citing open discussion, a safe and/or welcoming environment, and a clear explanation of how the data will be used as the main reasons for comfort. Several participants expressed value in creating dedicated space for older adults to discuss age-specific aspects of Type 1 diabetes management: “There is great value in listening to the not so good outcomes as we old ducks struggle with the technology…. How best to supply thoughtful, personal support to overcome hesitancy and get folks to actually embrace the technology [Workshop 1, Participant 2].” One participant additionally shared with the research team, “Having the experiences of others was most helpful. Being diabetic sometimes makes you feel alone. Getting to share with your age group was therapeutic [Workshop 3, Participant 4].” Other participant feedback on the group model building workshop is summarized in **Table 2**, in which participants indicated they liked sharing information and learning from others, finding peer support through shared experiences, the structure and pace of the workshop including small group sessions, age-specific discussions, the systems thinking framework, and the opportunity to contribute to research. Key aspects of feedback for future sessions focused on the duration of the session (where some participants indicated they would like a longer session and other indicated preference for a shorter session) and the lack of food or beverages provided by the research team.

**Table 2.**
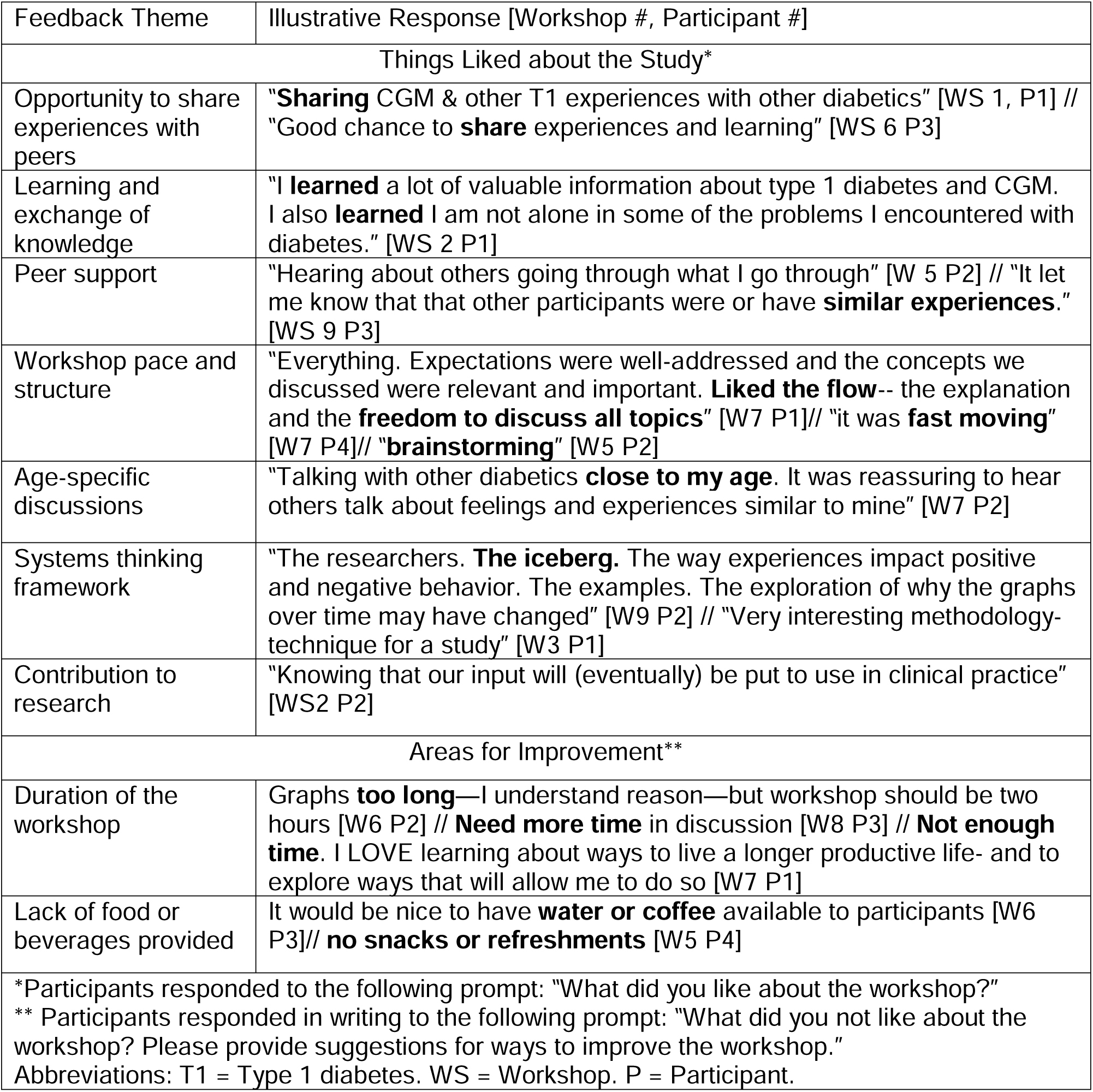
Illustrative feedback on the in-person group model building workshop from older adults living with Type 1 diabetes and their caregivers.

### Focus Group Discussion

The four focus group questions provided an opportunity to collect any last feedback or perspectives from study participants, but in general, did not reveal new themes or dynamics underlying CGM use that had not been identified through the preceding GMB activities. Participants expressed the sentiment that they had already shared their material they felt to be important relating to CGM use and non-use in older adults.

### Validation Interviews

27 of the 33 in-person study participants indicated that they were interested in providing feedback on the diagram. Following completion of the causal loop diagram, eight virtual validation interviews were conducted between March and April 2022. Each interview focused on validating a specific component of the diagram, including the core structure and the key feedback loops. Revisions made in validation were minor (i.e., modifications to variable names and addition of missing components) and included 1) connecting reactions from caregivers to a perceived sense of intrusiveness, an additional variable to indicate that alarm fatigue would be driven by frequency of alarms, and clarifying that improved HbA1c is associated with a sense of prolonging life alongside preventing complications, both of which promote future CGM use.

## Discussion

We applied group model building, a participatory approach to system dynamics modeling, to collect data from older adults with Type 1 diabetes and their caregivers through group workshops and individual validation interviews. This novel method of qualitative inquiry allowed us to diagram the structure of the ‘system’ of the multiple, interconnected variables that interrelate to produce four common emergent profiles of older-adult experiences using CGM (26, 27).

To our knowledge, few studies to date have extended participatory systems science methods to engage older adult patients. Thus, we consider our study to be a substantial contribution to the literature in that it demonstrates feasibility, acceptability, and value of this methodology to engage older adult patients and stakeholders in research relevant to their health and well-being. It is unclear why there are so few other studies to employ this method among older adults. One possibility is that the cognitive complexity of group model building may not be seen as feasible within older adults. A recent editorial in the *Journal of the American Medical Association* stated that “structural, or institutional, ageism is not only one of the most potent forms of bias that exists today, but also one of the least acknowledged ”(39). The authors go on to cite a report from the World Health Organization that underscores the long-standing history of institutional agism and the ways in which agism has become normalized across many domains both in and out of healthcare (40). A key finding of our study was that the didactic component of the study was well-received, and participants expressed positive feedback for the systems thinking framework, with more than 80% indicating interest in further engagement by feedback on the causal loop diagram. By contrast, the most challenging aspect of the study involved strategies to encourage drawing of behavior over time graphs to describe personal experiences with CGM. Future studies that apply group model building with older adult stakeholders will continue to shed insight into how the methodology can be made most accessible and best leveraged to elevate older adult voices as part of the clinical literature that informs future interventions for care and self-management.

Several aspects of the group model building study proved to be effective. The single-session, three-hour long workshop was associated with efficient recruitment and high attendance rates. Although we explored group model building in varied group sizes, we found that data collection was optimized at a group size of between four and six participants, as this size allowed for sufficient exchange of ideas but provided enough time for each participant to speak. Presenting an example of the systems thinking approach as part of the didactic component helped to solidify the framework and prepare participants for the activities to come. We intentionally selected to present an example focused around a potentially stigmatizing lifestyle change— increasing physical activity—to implicitly reinforce the value of group model building for diving deeper into clinical outcomes or trajectories that may be similarly stigmatized in clinical settings. We hoped that by showing the complexity underlying individual health-related decision-making, that participants would feel comfortable exploring the deeper trends that led to “less than ideal” CGM monitoring. We also drew and provided reference modes for the study, rather than asking study participants to brainstorm and co-create them. In the context of our research question, as well as the time-constraints and need to avoid participant fatigue, we found this approach to be effective. Further, personalizing the reference modes through named older adult characters helped to bridge the graphs to storytelling as part of the collective annotation.

A powerful aspect of the in-person workshops involved the ways in which participants and groups bonded over the duration of the study. We found that bonding occurred regardless of differences in demographics or clinical histories and was largely driven by resonance in day-to-day experiences in managing Type 1 diabetes or age-specific changes in older adulthood. The value of this peer support was reflected in study feedback. From a study design perspective, allowing time and space for long-form introductions prior to the structured presentations and activities was critical. Participants were generally eager to exchange tips or resources for Type 1 diabetes management and effective CGM use, although the research team always stressed the importance of talking with a healthcare provider about any clinical changes to their healthcare plan. Several participants expressed interest in continuing to meet with other participants in their workshops to continue conversations and the exchange of information and experiences.

There were several aspects of the study that were more challenging. One of the major limitations was the ability to recruit an adequately diverse sample to reflect the heterogeneity of the population of older adults living with Type 1 diabetes. The limited number of participants from underrepresented racial and ethnic groups (i.e. those of Black or African American race) and non-CGM users likely reflects a combination of selection bias related to recruiting from a single academic medical center, as well as a degree of survivor bias in which individuals with diabetes who did not have access to high-quality treatment or the ability to self-manage effectively earlier in disease duration were not represented; in cohort studies, excess mortality associated with Type 1 diabetes has been shown to disproportionately affect African American individuals compared to their Caucasian counterparts (5). Given the in-person component, the majority of study participants were also local to the city in which the academic medical center was located. Further, the selection bias associated with the COVID-19 vaccination requirement likely represents a complex medical and social bias, the effect of which on the study findings is difficult to characterize. Future work to include more diverse participants within group sessions is critically needed to ensure that conceptual models and diagrams are valid for this population.

There are other limitations to external generalizability of the findings. Our study did not include older adults with cognitive, visual, or hearing impairments, and thus cannot be generalized to all older adults. Because group model-building focuses on working together to describe stories, having a group that shares the same language is also a requirement, further narrowing our sample and thus the overall representativeness of this study.

Future group model building studies for older adults may consider the following points of learning. First, data collection may be enrichened with regards to underrepresented views, including one or more group study sessions dedicated to capturing the experiences of such individuals, without dilution or bias from influence of subgroups that tend to be over-represented (41). In our study, a small proportion of older adults living with Type 1 diabetes elected to bring a caregiver with them to the study. Future studies which aim to integrate diverse stakeholder perspectives, such as those from caregivers, may consider recruiting these stakeholders independently for participation in a focused workshop of caregiver participants only. Although we included a brief focus group discussion as part of our workshop, we found that participant responses were very brief or largely redundant with data collected through the drawing and annotation exercises; thus, this section of the study did not significantly expound upon or enrich data. We believe this reflects, in large part, the open-ended nature of the preceding group model building activities and vigorous group discussion. Finally, in review of the transcripts of the study, we found that participant narratives yielded significant contextual information, personal narrative, or other ‘foreground’ for clinical questions that are better captured and described by inductive qualitative analysis techniques. Our research team aims to explore how the results from system dynamics analyses and varied qualitative analyses (42) can be triangulated to provide a complementary, comprehensive view of the lived experiences of older adults with Type 1 diabetes, as well as their complex experiences with technology.

There are several questions remaining. It remains unclear how providers can be integrated into group model building, and if the potential for power dynamics between patient and provider stakeholders may influence the quality of group model building in mixed groups. Recruiting patient and provider stakeholders across institutions or holding provider-specific workshops may avoid this problem, although validation between groups may prove more challenging for the latter. It is also unknown how the causal loop diagrams may change if older adult participants are directly involved in the diagraming process, and how the diagrams would change between sessions. Studies that included older adult participants in direct diagraming may need to explore various study formats to ensure that burden remains low, and participants do not experience significant fatigue. Given that the number of feedback loops in our causal loop diagram exceeded 100, it is likely that generating comprehensive diagrams with participants directly may involve more than one workshop, thus increasing the burden of the research study and potentially hampering recruitment outcomes.

It has been argued that systems thinking and systems science methods, such as system dynamics, remain underutilized for complex health problems (26, 27, 43, 44). For the study objective, which demands an awareness of more complexity than traditional research methods (27), group model building provided a novel approach among older adults to comprehensively investigate and describe multiple, interrelated factors, as well as their complex interactions over time. The holistic view of experiences as the behavior of a complex system offers the opportunity to not only describe, but start to untangle the mechanisms that shape older adults’ experiences with Type 1 diabetes technology and self-management (35, 38). In future work, our team aims to use the causal loop diagrams to identify outcome sets which represent ‘suboptimal CGM responses’ that signal the need for additional resources, education, or support, as well as the system structure of the factors that interact to produce each response; we will use this problem definition as the basis for efforts to develop new strategies to address and prevent suboptimal trajectories associated with CGM use in older adults with Type 1 diabetes. We are also enthusiastic to extend the group model building methodology, and integrate lessons learned in the present study, to continue to engage older adults as primary stakeholders in research to promote longevity and healthy aging across a range of clinical contexts.

## Supporting information

The example and didactic language are presented in full in Appendix A: Primer to Systems Thinking and Systems Mapping.

A sample workshop packet from the latter five workshops can be found in Appendix B: Group Model Building Workshop Packet.

## Data Availability

All data produced in the present study are available upon reasonable request to the authors.

## Acknowledgements

We are grateful to the participants of the CGM Older Adult Stakeholder Mapping Workshop research study. We are grateful to Noelé Daniels for her assistance with the study space and scheduling procedures. Damilola Ayinde, Kabir Dewan, Maya Loga, and Sharita Thomas provided research assistance during the workshops.

## Supporting Information Captions

Appendix A: Primer to Systems Thinking and Systems Mapping

Appendix B: Group Model Building Workshop Packet.

## References

1. Atkinson MA, Eisenbarth GS, Michels AW. Type 1 diabetes. The Lancet. 2014;383(9911):69–82.

2. Holt RI, DeVries JH, Hess-Fischl A, Hirsch IB, Kirkman MS, Klupa T, et al. The management of type 1 diabetes in adults. A consensus report by the American Diabetes Association (ADA) and the European Association for the Study of Diabetes (EASD). Diabetes Care. 2021;44(11):2589–625.

3. Kirkman MS, Briscoe VJ, Clark N, Florez H, Haas LB, Halter JB, et al. Diabetes in older adults. Diabetes care. 2012;35(12):2650–64.

4. Munshi MN, Meneilly GS, Rodríguez-Mañas L, Close KL, Conlin PR, Cukierman-Yaffe T, et al. Diabetes in ageing: pathways for developing the evidence base for clinical guidance. The Lancet Diabetes & Endocrinology. 2020;8(10):855–67.

5. Secrest AM, Becker DJ, Kelsey SF, LaPorte RE, Orchard TJ. Cause-specific mortality trends in a large population-based cohort with long-standing childhood-onset type 1 diabetes. Diabetes. 2010;59(12):3216–22.

6. Association AD. 12. Older adults: standards of medical care in diabetes—2019. Diabetes Care. 2019;42(Supplement 1):S139-S47.

7. Sue Kirkman M, Briscoe VJ, Clark N, Florez H, Haas LB, Halter JB, et al. Diabetes in older adults: a consensus report. Journal of the American Geriatrics Society. 2012;60(12):2342–56.

8. Kirkman MS, Briscoe VJ, Clark N, Florez H, Haas LB, Halter JB, et al. Diabetes in older adults: consensus report. Journal of the American Geriatrics Society. 2012;60(12):2342.

9. LeRoith D, Biessels GJ, Braithwaite SS, Casanueva FF, Draznin B, Halter JB, et al. Treatment of diabetes in older adults: an Endocrine Society clinical practice guideline. The Journal of Clinical Endocrinology & Metabolism. 2019;104(5):1520–74.

10. Committee ADAPP, Committee: ADAPP. 13. Older Adults: Standards of Medical Care in Diabetes—2022. Diabetes Care. 2022;45(Supplement_1):S195–S207.

11. Klonoff DC. Continuous glucose monitoring: roadmap for 21st century diabetes therapy. Diabetes care. 2005;28(5):1231–9.

12. Committee ADAPP, Committee: ADAPP. 7. Diabetes Technology: Standards of Medical Care in Diabetes—2022. Diabetes Care. 2022;45(Supplement_1):S97–S112.

13. Pratley RE, Kanapka LG, Rickels MR, Ahmann A, Aleppo G, Beck R, et al. Effect of continuous glucose monitoring on hypoglycemia in older adults with type 1 diabetes: a randomized clinical trial. Jama. 2020;323(23):2397–406.

14. Munshi M, Slyne C, Davis DQ, Michals A, Sifre K, Dewar R, et al. Use of Technology in Older Adults with Type 1 Diabetes: Clinical Characteristics and Glycemic Metrics. Diabetes technology & therapeutics. 2022;24(1):1–9.

15. Ruedy KJ, Parkin CG, Riddlesworth TD, Graham C. Continuous glucose monitoring in older adults with type 1 and type 2 diabetes using multiple daily injections of insulin: results from the DIAMOND trial. Journal of diabetes science and technology. 2017;11(6):1138–46.

16. Polonsky WH, Peters AL, Hessler D. The impact of real-time continuous glucose monitoring in patients 65 years and older. Journal of diabetes science and technology. 2016;10(4):892–7.

17. Divan V, Greenfield M, Morley CP, Weinstock RS. Perceived burdens and benefits associated with continuous glucose monitor use in type 1 diabetes across the lifespan. Journal of Diabetes Science and Technology. 2022;16(1):88–96.

18. Gubitosi-Klug RA, Braffett BH, Bebu I, Johnson ML, Farrell K, Kenny D, et al. Continuous Glucose Monitoring in Adults With Type 1 Diabetes With 35 Years Duration From the DCCT/EDIC Study. Diabetes Care. 2022.

19. Krishnaswami A, Beavers C, Dorsch MP, Dodson JA, Masterson Creber R, Kitsiou S, et al. Gerotechnology for older adults with cardiovascular diseases: JACC state-of-the-art review. Journal of the American College of Cardiology. 2020;76(22):2650–70.

20. Glass TA, McAtee MJ. Behavioral science at the crossroads in public health: extending horizons, envisioning the future. Social science & medicine. 2006;62(7):1650–71.

21. Litchman ML, Allen NA. Real-time continuous glucose monitoring facilitates feelings of safety in older adults with type 1 diabetes: a qualitative study. Journal of diabetes science and technology. 2017;11(5):988–95.

22. Kerr D, Glantz N. Diabetes, like COVID-19, is a wicked problem. The Lancet Diabetes & Endocrinology. 2020.

23. Homer JB, Hirsch GB. System dynamics modeling for public health: background and opportunities. American journal of public health. 2006;96(3):452–8.

24. Lich KH, Ginexi EM, Osgood ND, Mabry PL. A call to address complexity in prevention science research. Prevention science. 2013;14(3):279–89.

25. Cilenti D, Issel M, Wells R, Link S, Lich KH. System dynamics approaches and collective action for community health: An integrative review. American Journal of Community Psychology. 2019;63(3-4):527–45.

26. Apostolopoulos Y, Lich KH, Lemke MK. Complex Systems and Population Health: A Primer: Oxford University Press; 2019.

27. Luke DA, Stamatakis KA. Systems science methods in public health: dynamics, networks, and agents. Annual review of public health. 2012;33:357–76.

28. Meadows DH. Thinking in systems: A primer: chelsea green publishing; 2008.

29. Zavacki JF. Group model building: facilitating team learning using system dynamics. Quality Progress. 1997;30(11):124.

30. Hovmand PS, Andersen DF, Rouwette E, Richardson GP, Rux K, Calhoun A. Group model-building ‘scripts’ as a collaborative planning tool. Systems Research and Behavioral Science. 2012;29(2):179–93.

31. Apostolopoulos Y, Lemke MK, Barry AE, Lich KH. Moving alcohol prevention research forward—Part I: Introducing a complex systems paradigm. Addiction. 2018;113(2):353–62.

32. Kim DH. Introduction to systems thinking: Pegasus Communications Waltham, MA; 1999.

33. Monat JP, Gannon TF. What is systems thinking? A review of selected literature plus recommendations. American Journal of Systems Science. 2015;4(1):11–26.

34. Andersen DF, Richardson GP. Scripts for group model building. System Dynamics Review: The Journal of the System Dynamics Society. 1997;13(2):107–29.

35. Sterman JD. Learning from evidence in a complex world. American journal of public health. 2006;96(3):505–14.

36. Bayer S. Business dynamics: Systems thinking and modeling for a complex world. JSTOR; 2004.

37. Naumann RB, Kuhlberg J, Sandt L, Heiny S, Apostolopoulos Y, Marshall SW, et al. Integrating complex systems science into road safety research and practice, part 1: review of formative concepts. Injury prevention. 2020;26(2):177–83.

38. Apostolopoulos Y, Lich KH, Lemke MK. Complex Systems and Population Health: Oxford University Press; 2020.

39. Levy BR. The Role of Structural Ageism in Age Beliefs and Health of Older Persons. JAMA Network Open. 2022;5(2):e2147802–e.

40. WHO. Global Report on Ageism. World Health Organization Geneva, Switzerland; 2021.

41. LaVaccare S, Diamant AL, Friedman J, Singh KT, Baker JA, Rodriguez TA, et al. Healthcare experiences of underrepresented lesbian and bisexual women: a focus group qualitative study. Health Equity. 2018;2(1):131–8.

42. Saldaña J. Coding and analysis strategies. The Oxford handbook of qualitative research 2014.

43. Trochim WM, Cabrera DA, Milstein B, Gallagher RS, Leischow SJ. Practical challenges of systems thinking and modeling in public health. American journal of public health. 2006;96(3):538–46.

44. Van Wave TW, Scutchfield FD, Honoré PA. Recent advances in public health systems research in the United States. Annual review of public health. 2010;31:283–95.

