## Supplementary material for "Use of Systems Thinking and Group Model Building Methods to Understand Patterns of Continuous Glucose Monitoring Use Among Older Adults with Type 1 Diabetes": The example and didactic language are presented in full in Appendix A: Primer to Systems Thinking and Systems Mapping.

Appendix A: Primer to Systems Thinking and Systems Mapping Provided to Study Participants as Optional Take Home Material

### CGM Older Adult Stakeholder Mapping Workshop

#### Introduction to Systems Thinking and System Mapping

*This is a resource to overview what systems thinking means and how we can use system mapping to better understand the complex factors that influence health and wellness.*

**Table of Contents**

1. Introduction to Systems Thinking

2. An Example of Using Systems Thinking: Betty and Jan’s New Year’s Resolution

   *The Shared Resolution: A Daily Step Goal of 10,000 Steps*

   *Different Outcomes of the Same Resolution*

   *Behavior Over Time Graphs*

   *Related Trends*

   *System Maps*

   *Improving System Maps with New Information*

3. How we can use systems thinking and system maps to better support patients achieving health and wellness

#### 1. Introduction to Systems Thinking

We live in a complex world and staying healthy is a complex task.

There are lots of different things, or factors, that influence someone's overall health and wellness. These factors are often related to each other, and those relationships change over time in unexpected ways. Further, the ways in which they may be related or change over time may be different for different people. For these reasons, it is often difficult to understand how and why someone may do certain things, practice certain habits, or experience certain patterns in their health.

Systems thinking is an approach that aims to sort out these complex relationships, and better understand how and why things happen. A good metaphor for systems thinking is an iceberg.

Only the top of an iceberg may be visible above the waterline.

Although this is the part that we actually "see," it is actually only a very small part of the full iceberg. We can't really understand the depth or shape of the iceberg without diving underwater to see the full picture.

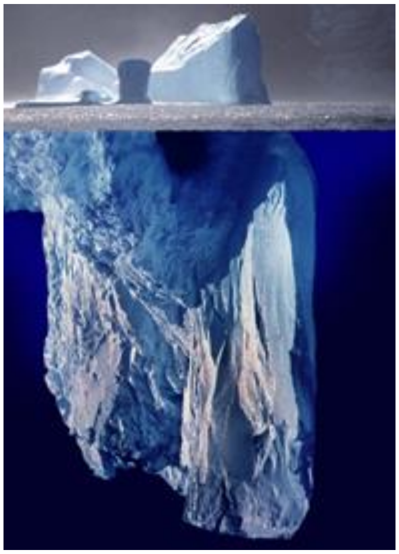

*The iceberg image is work by Uwe Kils http://www.ecoscope.com/iceberg/. Creative Commons CC BY-SA 3.0*

We can think about health outcomes or events in a similar way. We often focus on single outcomes or events that we observe, measure, or report in the doctor's office. (This is the tip of the iceberg). However, staying healthy requires us to do things both to manage prevent diseases (exercising, eating healthfully, etc.) and manage diseases (taking medications, monitoring or measuring health outcomes at home, etc.). In order to understand why an outcome or event happened, we need to look 'below the waterline' to understand what other factors that contributed and how, the relationships between those factors, and the underlying beliefs that individuals, their friends and family, and their care providers may have. We can break those out into three levels of understanding that are all below the waterline: Patterns of Behavior, System Structure, and Goals & Values.

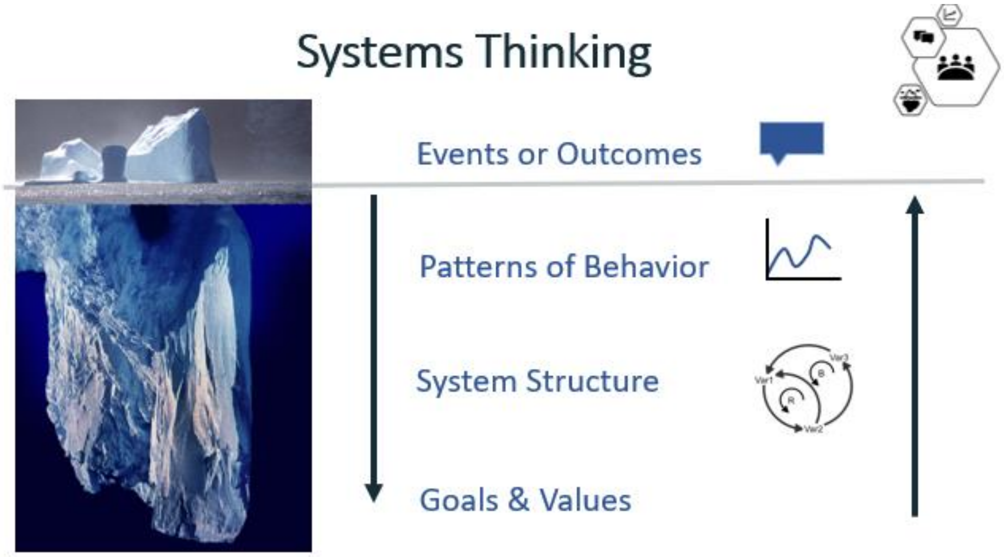


*The iceberg image is work by Uwe Kils http://www.ecoscope.com/iceberg/. Creative Commons CC BY-SA 3.0*

We can make those levels more specific to health and wellness.

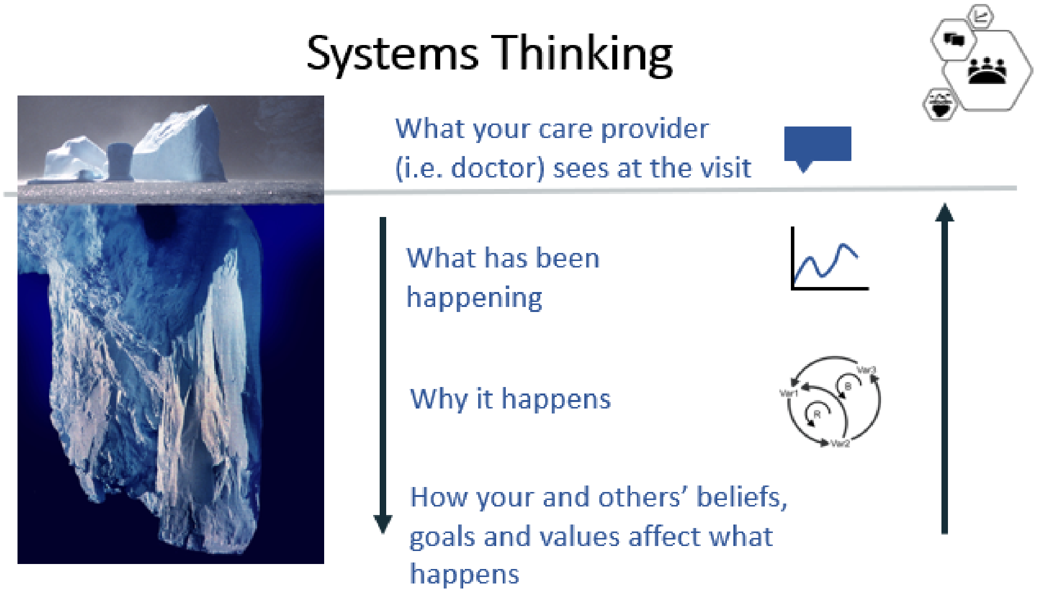


*The iceberg image is work by Uwe Kils http://www.ecoscope.com/iceberg/. Creative Commons CC BY-SA 3.0*

An important tool for systems thinking, and specifically to dive below the waterline, is drawing picture. We use simple graphs called 'behavior over time graphs' to think about how and why events happen by drawing the patterns or changes in other factors.

We can use these graphs to describe the main problem that we care about, as well as all of the related trends.

Here is an example of different behavior of time graphs.

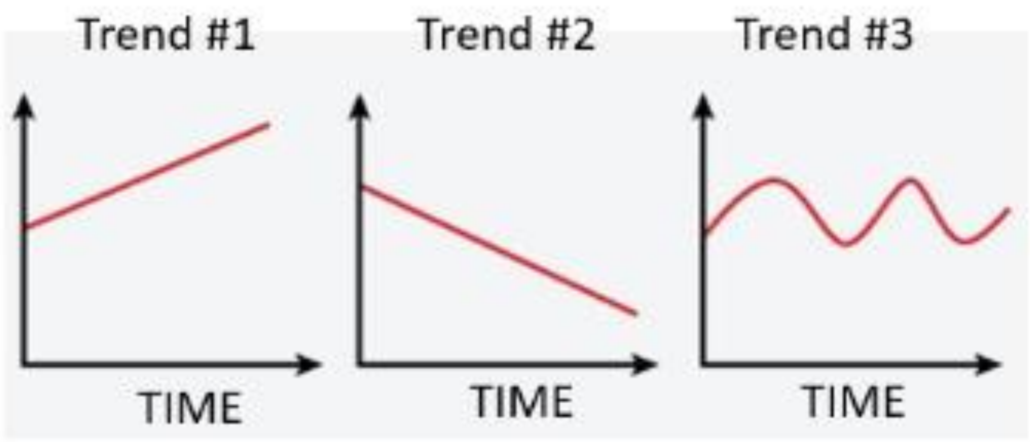

Once we have drawn graphs of the main problem we care about, and the related trends that we believe are most important, we can use those graphs to put all that information together in one drawing. This drawing is called a system map, and it helps us to understand how each trend is related to other trends, as well as the main problem we are trying to solve. We will walk through each of these steps in an example.

#### 2. *An Example of Using Systems Thinking: Betty and Jan's New Year's Resolution*

Let's work through an example of systems thinking. The example will show how we can use systems thinking to better understand the complexity of a seemingly simple behavior for staying healthy: daily exercise. We will also show how taking the time to dive below the waterline can reveal opportunities to better support and health people achieve their best possible health and wellness.

*The Shared Resolution: A Daily Step Goal of 10,000 Steps*

Consider two characters: Betty and Jan are both 74-year-old women who have been friends for a long time. They live in the same town and often meet for lunch to catch up on each other's lives.

As the new year approaches, Betty and Jan have been discussing their New Year's Resolution for the upcoming year. They both agree that they would like to exercise more. Specifically, they both aim to walk 10,000 steps each day. They decide that they will both undertake this 'daily step goal' as their New Year's Resolution.

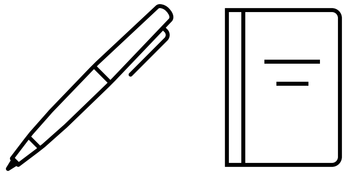

Resolution: "I will walk 10,000 steps every day"

*Different Outcomes of the Same Resolution*

Fast-forward to June: Betty is consistently walking 10,000 steps every day, but Jan is not walking at all.

Both Betty and Jan wanted to keep this resolution. What happened, and why? Let's work through the iceberg metaphor to try to understand. Here, the tip of the iceberg is the event or outcome that we observe in June, six months after setting the resolution.

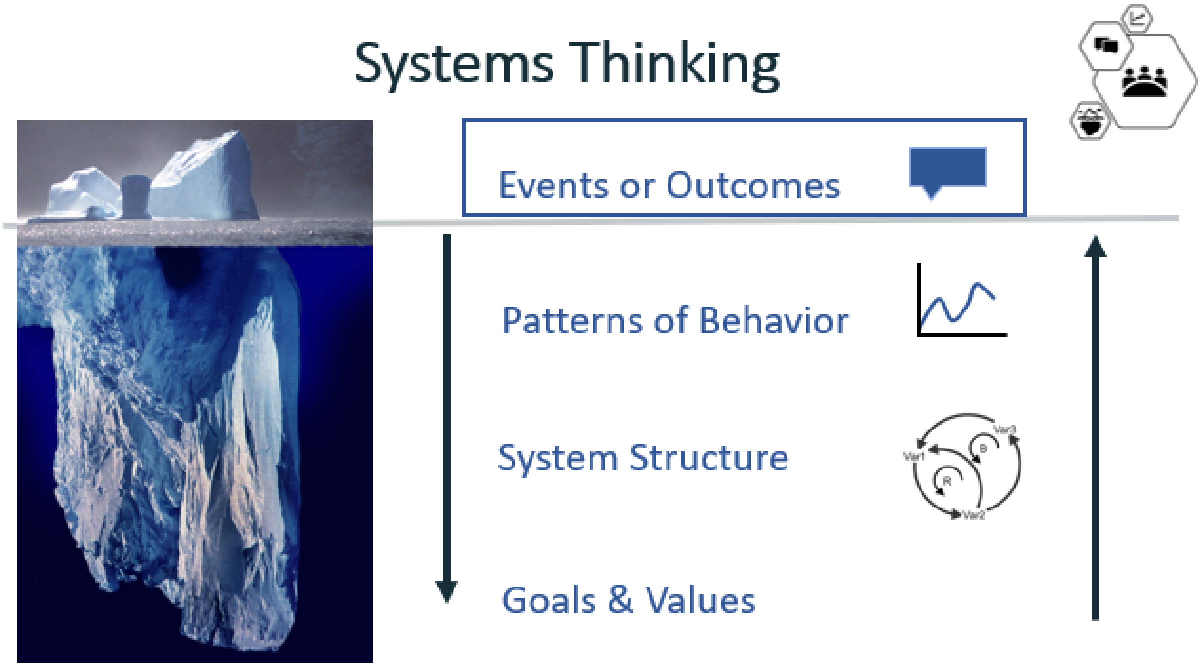


*The iceberg image is work by Uwe Kils http://www.ecoscope.com/iceberg/. Creative Commons CC BY-SA 3.0*

At that moment in time, Betty has been able to keep up with her resolution and is walking daily, but Jan has not.

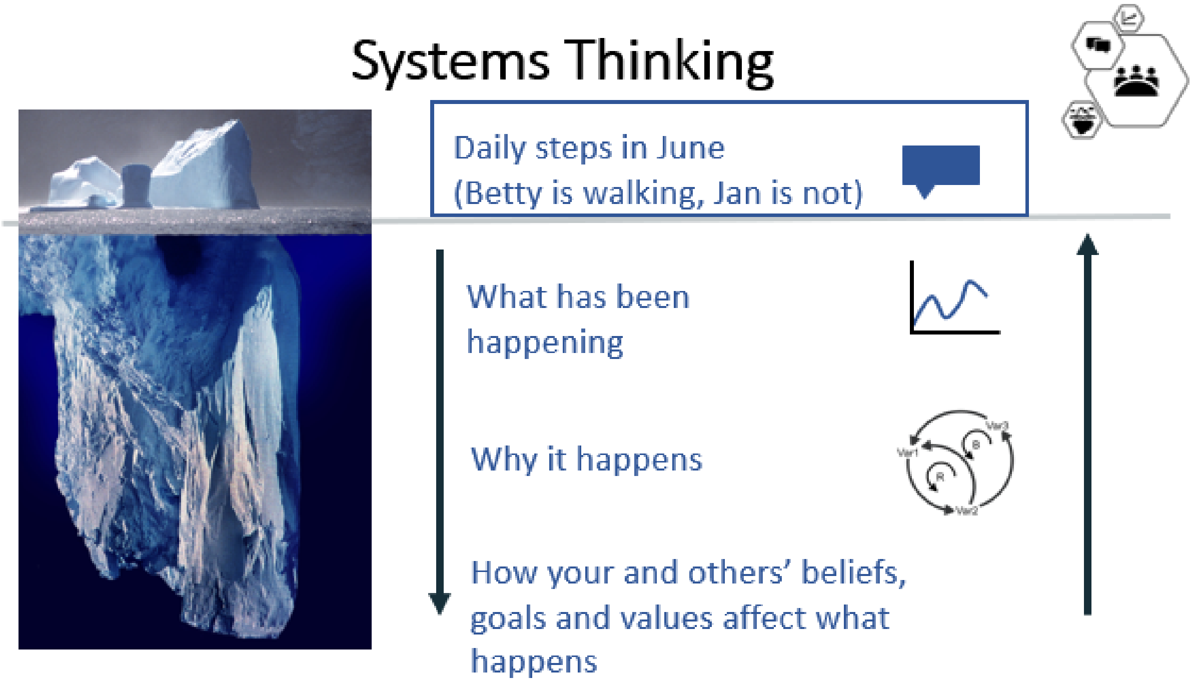


The iceberg image is work by Uwe Kils http://www.ecoscope.com/iceberg/. Creative Commons CC BY-SA 3.0

*Behavior Over Time Graphs*

Let's move below the waterline to see why there were different outcomes of the same resolution. Our main question now is, what happened?

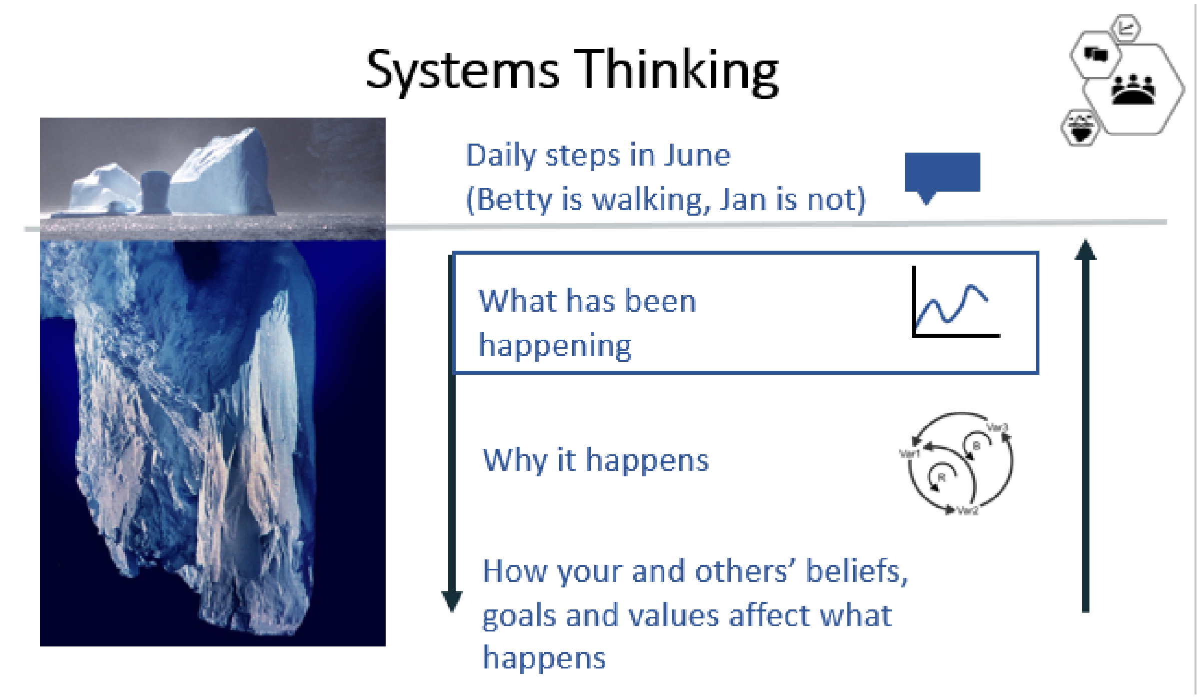


The iceberg image is work by Uwe Kils http://www.ecoscope.com/iceberg/. Creative Commons CC BY-SA 3.0

This is where we start to use behavior over time graphs. We can first look back to see how daily steps have changed in the months leading up to June.

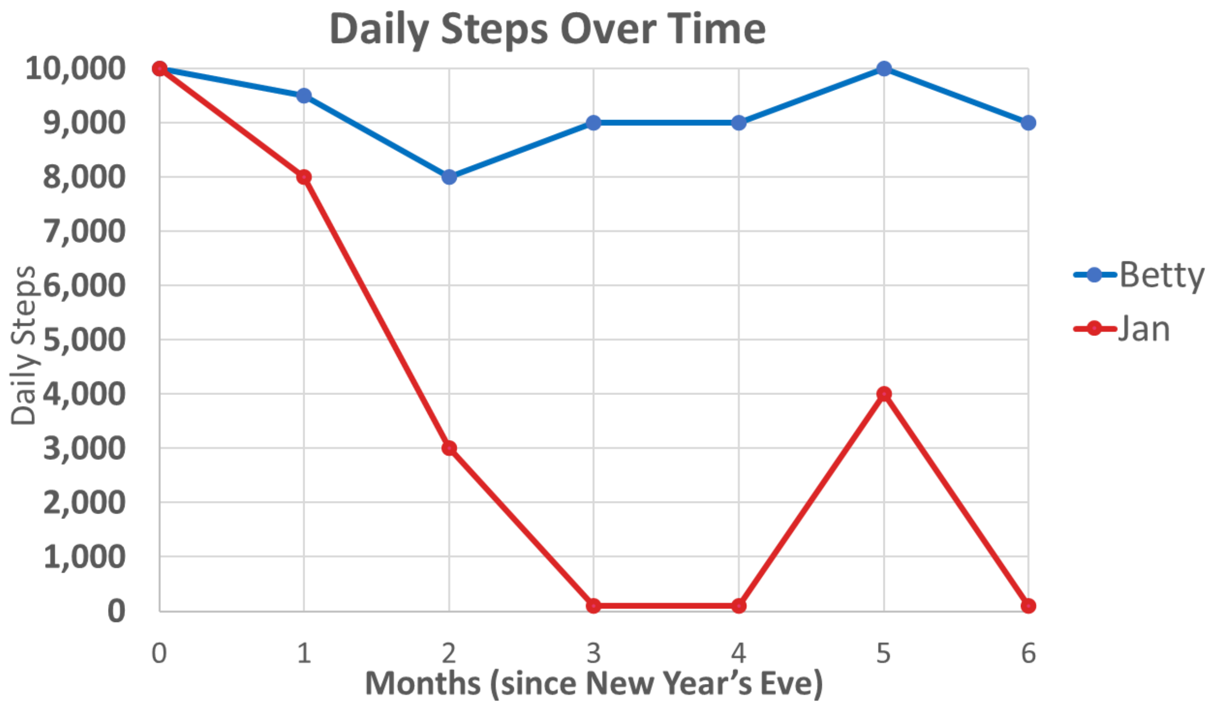


This graph shows us that not only were the outcomes at 6 months different, but Betty and Jan had very different experiences that led up to that outcome, in terms of their daily steps between December and June.

*Related Trends*

We can continue to move down the iceberg to ask, what other trends are related? When we think about related trends, there are no right or wrong answers.

Usually, we focus on nouns (e.g., people, places, things, ideas, etc.) and something that can increase or decrease over time. These factors do not need to have a formal scale, and they can be causes or consequences of the problem we are trying to understand.

Betty and Jan’s daily steps may have been related to how feasible they believed the daily step goal of 10,000 steps felt, their level of physical fitness, muscle soreness, or arthritis pain (knee pain, for example). Let’s think through these related trends for Betty, first.

Recall that this is how Betty's daily steps looked over time. In general, she walked between 8,000 and 10,000 steps most days since setting her resolutions.

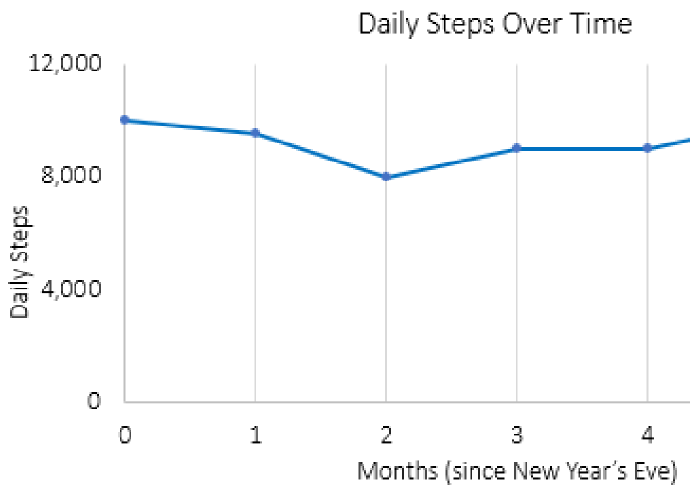


Here are her related trends. We can make notes to explain the related trends and why we draw them the way we do.

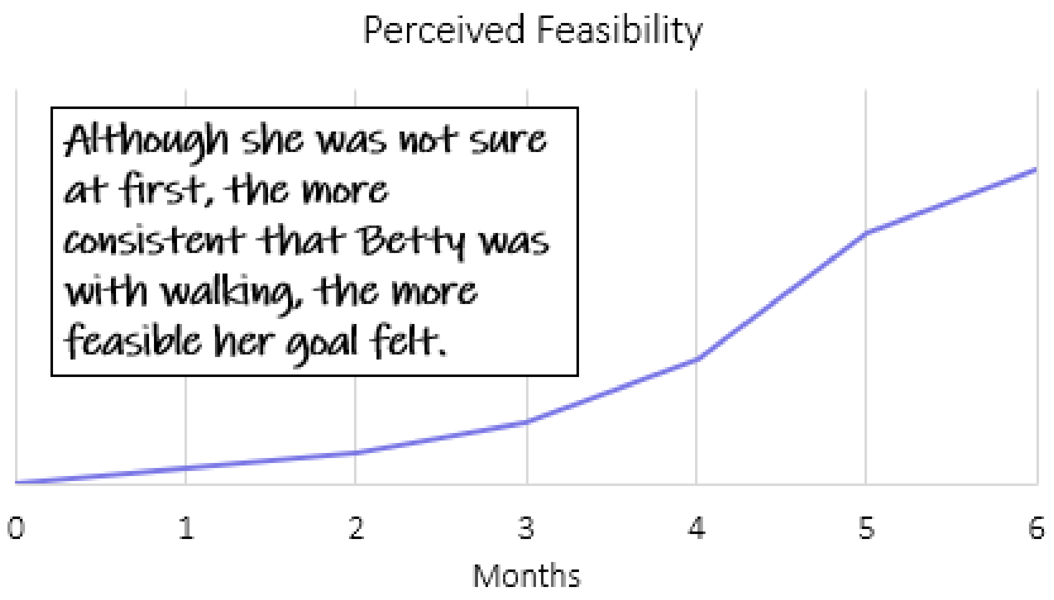

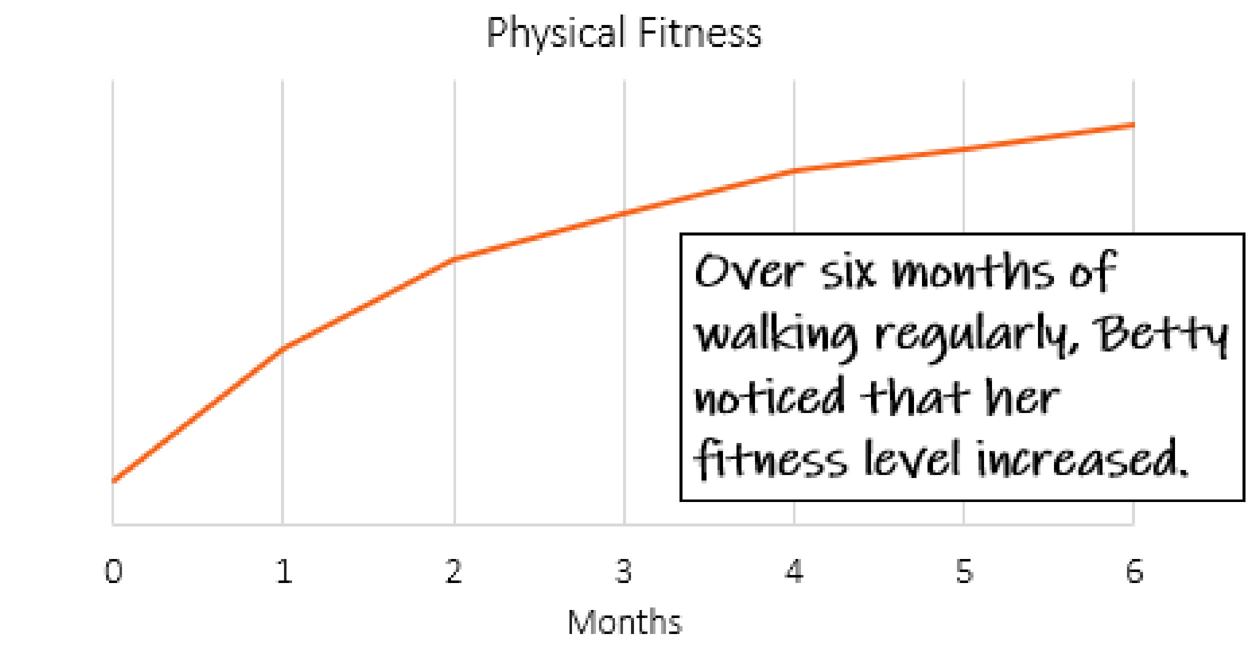
Physical Fitness
Over six months of walking regularly, Betty noticed that her fitness level increased.
0
1
2
3
4
5
6
Months

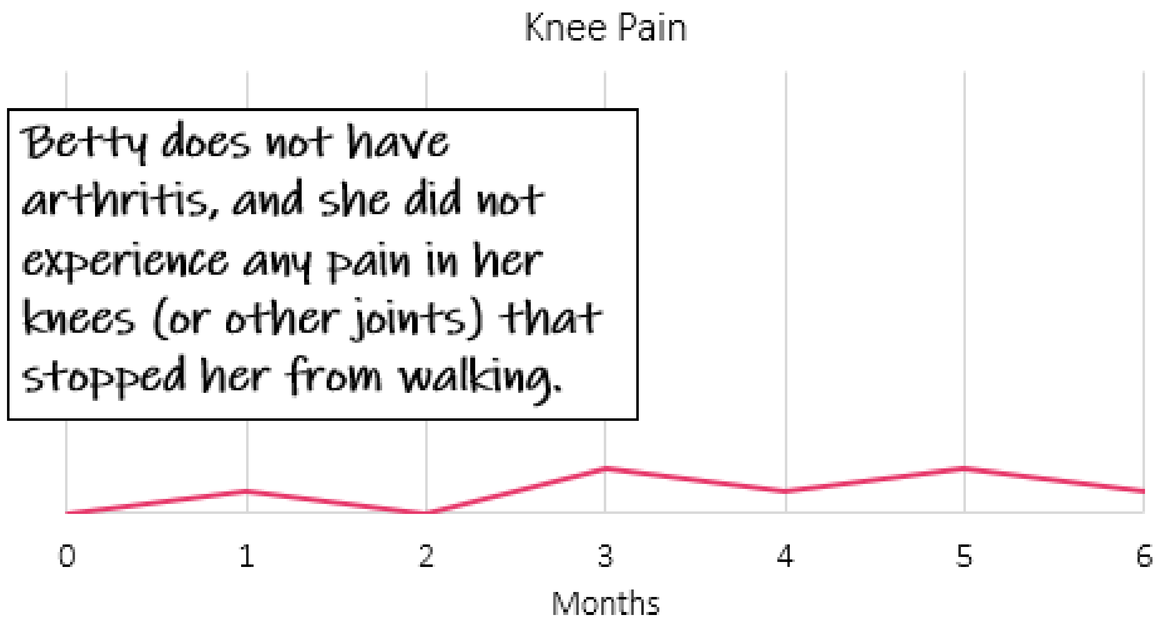
Knee Pain
Betty does not have arthritis, and she did not experience any pain in her knees (or other joints) that stopped her from walking.
0
1
2
3
4
5
6
Months

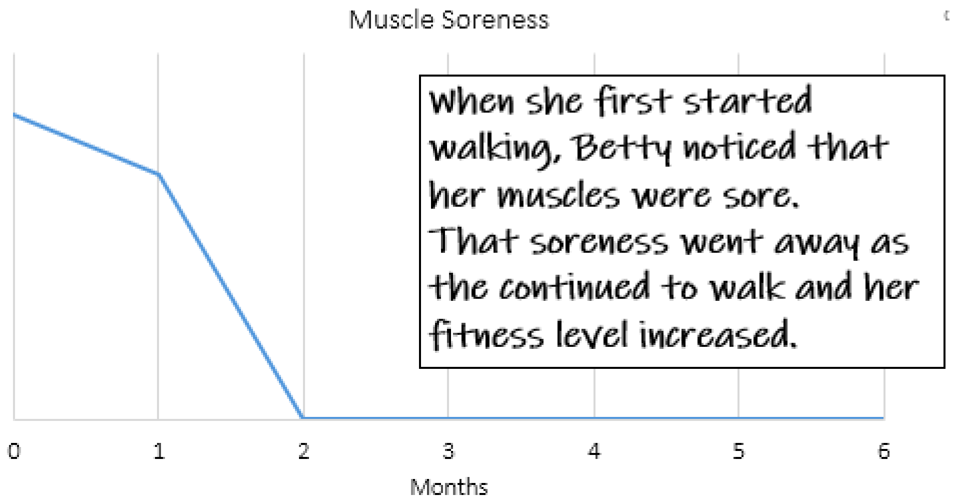


Other trends may include the time she spent with friends while walking, energy throughout the day, and how much she enjoyed walking.

Here is how it all looks together:

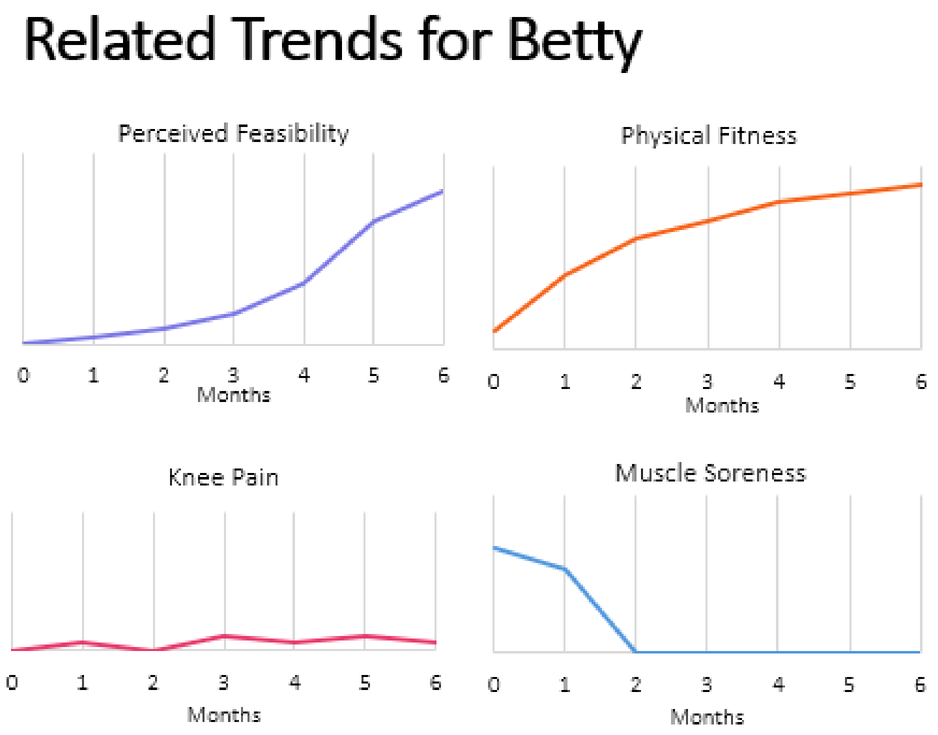

And, here is a story we could gather from these trends:

Betty started walking 10,000 steps her day shortly after the New Year. When she started walking, she had muscle soreness, but that went away as she continued to walk and her physical fitness increased.

The more the walked, and the better her physical fitness became, the more feasible her daily goal felt. She did not have pain that stopped her.

Now, let’s think through the same trends for Jan. Recall that this is how Jan’s daily steps looked over time. She started out strong, but her daily walking fell by March, and she although it looks like she tried to increase her daily steps, she was not able to do so.

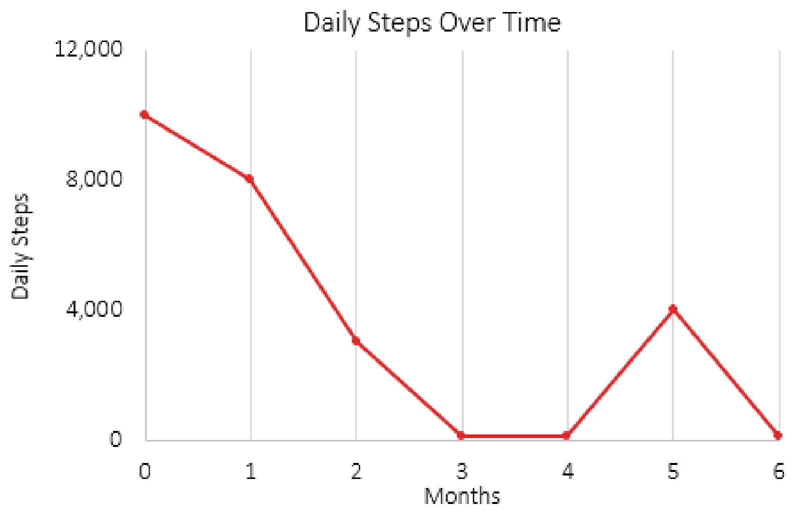


Here are her related trends:

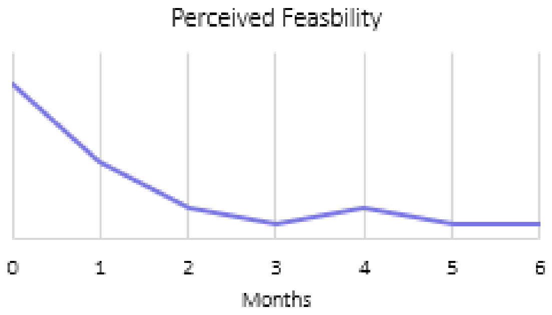


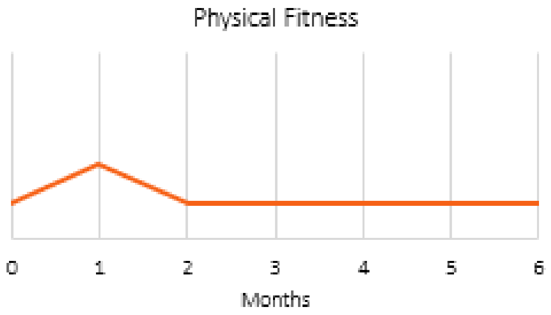


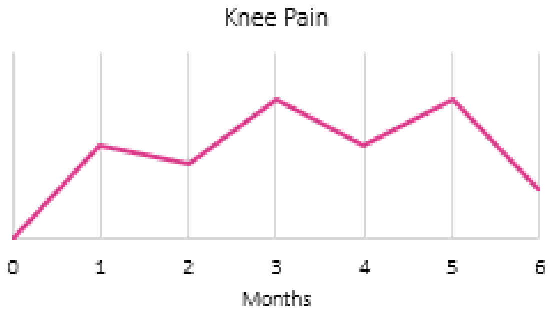


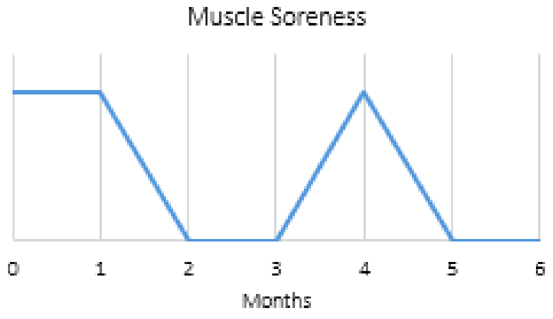

Here is the story we might be able to tell, based on these trends.

| |
|---|
| Jan was not able to keep walking because of muscle soreness that she experienced early on, as well as chronic knee pain from her arthritis.<br><br>We can see that when muscle soreness and knee pain increased, daily steps decreased. The rest helped the pain, but they recurred when she tried to walk again. |

*System Maps*

Let's move even farther below the waterline. Our main question now is, why did those trends happen?

To understand why, we are going to put together these trends as part of a larger "system" by drawing a 'map' of the trends to understand how they are related.

A system is a set of interconnected factors that interact to produce an effect that is greater or different than any one of its parts. This is called 'system-mapping,' and it is a way to sort out these complex relationships and better understand how and why things occur. For this part, we will combine what we know about Betty and Jan's experiences with their resolutions.

We start with our *factors*, or trends; these are the things that that can go up or down. We can start with any factor. Here, we will start with the number of days achieving the step goal, since this is an outcome we really care about. We can also add another important related trend, the perceived feasibility of the goal.

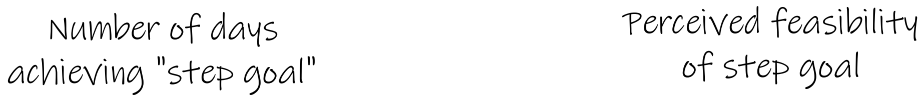


We will draw arrows that connect them. We label the arrows with a "+" because the variables are moving in the same direction. An increase in one results in an increase in another. Here, the more that someone achieves their daily step goal, the more feasible they will believe that goal to be for them.

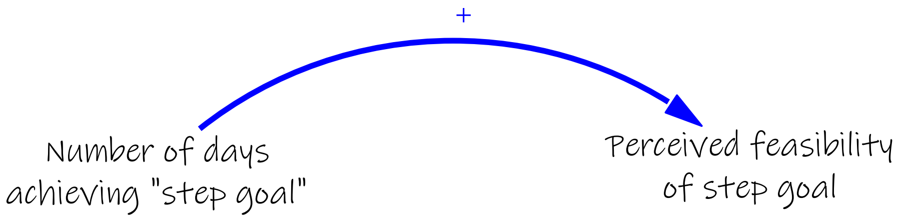


Continuing the story, the more feasible that a goal feels, the more likely someone may be to achieve it. To show this, we draw another "+" arrow back from the "perceived feasibility of step goal" factor to the "number of days achieving the step goal" factor.

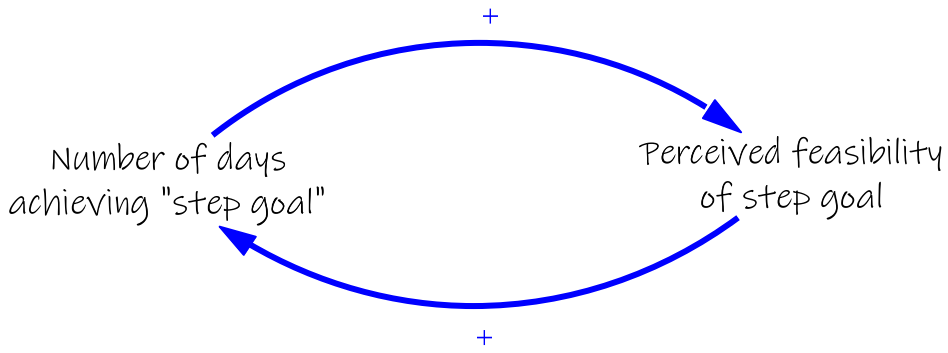


Notice here that we just created a closed, circular path. These circular paths create what we call reinforcing loops, where change in one variable add to the previous changes and keep the change going in the same direction. You can think of it as a 'snowballing' effect. Here, we started with an increase in the number of days achieving a step goal, which increased the perceived feasibility of that goal and came back around again to increase the number of days the goal was achieved.

We indicate that this is a reinforcing loop with an "R" on the map. We can label it to describe what is happening.

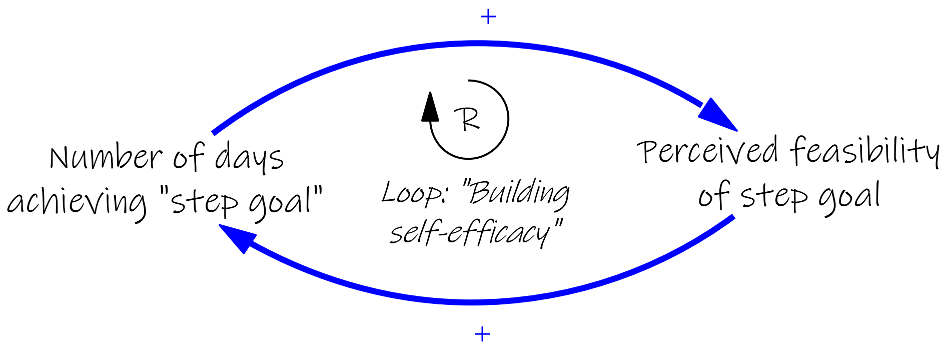


Let's add another trend that we know was important: muscle soreness.

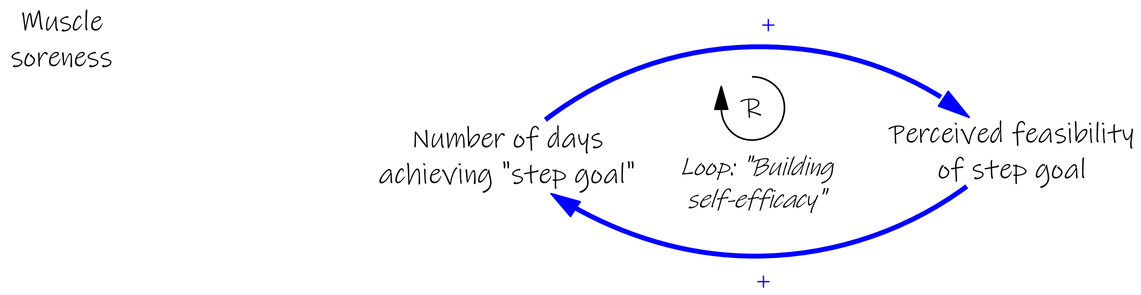


As Betty and Jan started walking, they experienced more muscle soreness, so we draw another “+” arrow.

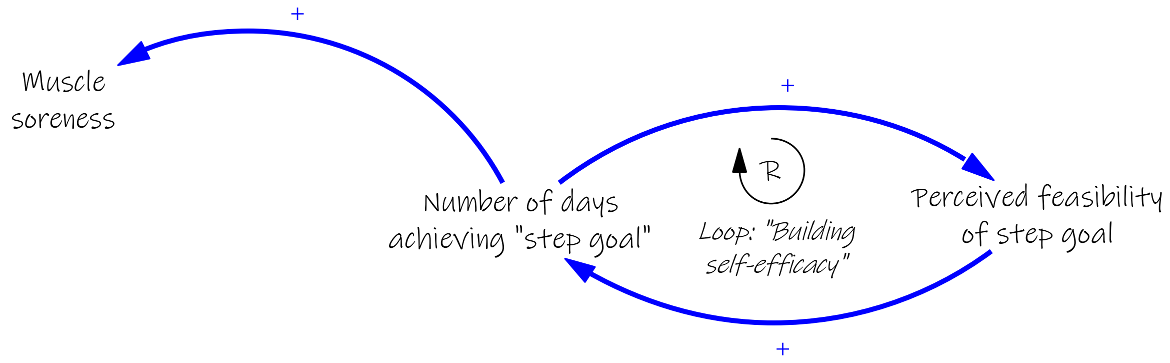


That soreness made the thought of going for a walk less desirable, so we draw an arrow and label it with a “-” to indicate that these factors move in opposite direction. An increase in one causes a decrease in the other.

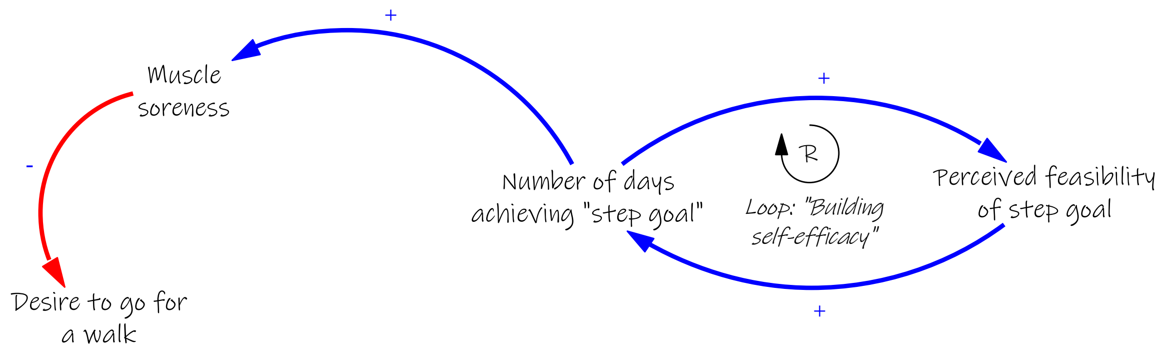


Let’s consider how desire to go for a walk is related to the number of days achieving a step goal. If walking is more desirable, someone may be more likely to do it. So, we use a “+” arrow to connect these factors.

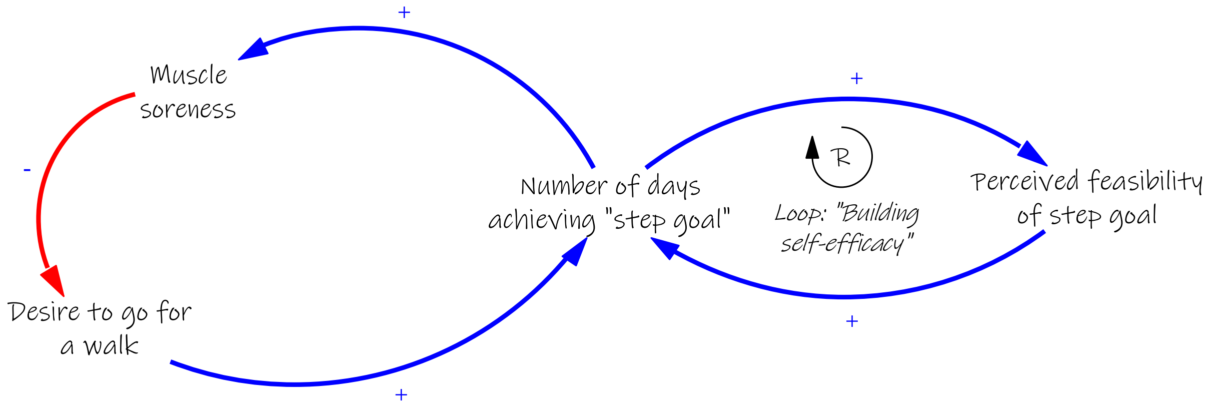

We just created another closed, circular path. But this one has a "-" inside of it. This is an example of a balancing loop. Balancing loops result when a change in one factor produces change in another variable in an opposite direction that ultimately limiting any further changes. You can think of this like a 'thermostat' effect. Here, an increase in days meeting the step goal increased muscle soreness, which in turn decreased the desire to go for a walk and likelihood of walking the next day.

We indicate that this is a balancing loop with an "B" on the map. We can label it to describe what is happening. Here, we call this "Post-walk soreness."

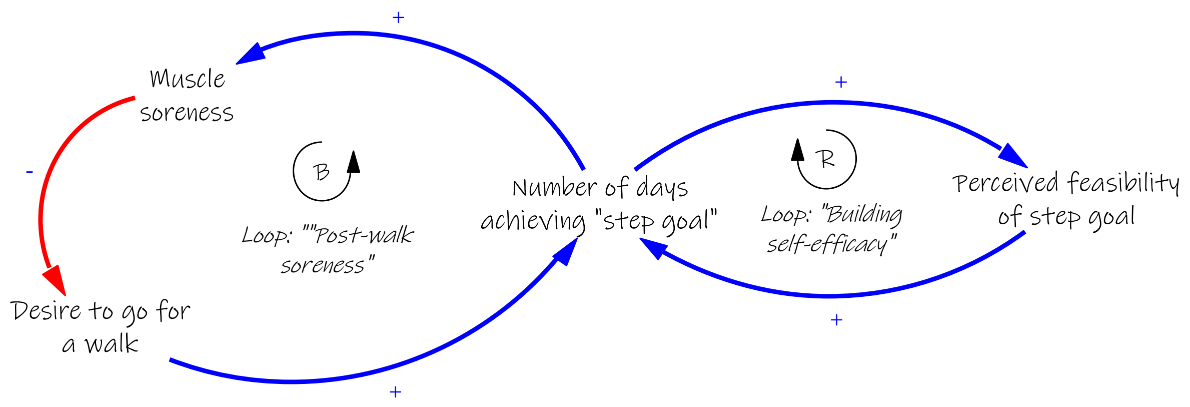


A simple rule for determining if a loop is reinforcing or balancing is to count the number of "-'s". Zero or even number of -'s = reinforcing loop. An odd number of -'s = balancing loop.

Understanding feedback loops is important because it explains why certain trends might happen. For example, if there are a lot of reinforcing loops, we might expect to see trends in our system that are increasing or decreasing very quickly. If there are more balancing loops, we might expect to see more examples of trends that start to change in one direction, but then that change slows down or is even stops all together. Most systems have a mix of reinforcing and balancing loops.

Another important thing we can show on our maps are time delays, or relationships between two factors that may play out over time versus immediately.

Consider physical fitness. As daily walking increases, physical fitness will increase.

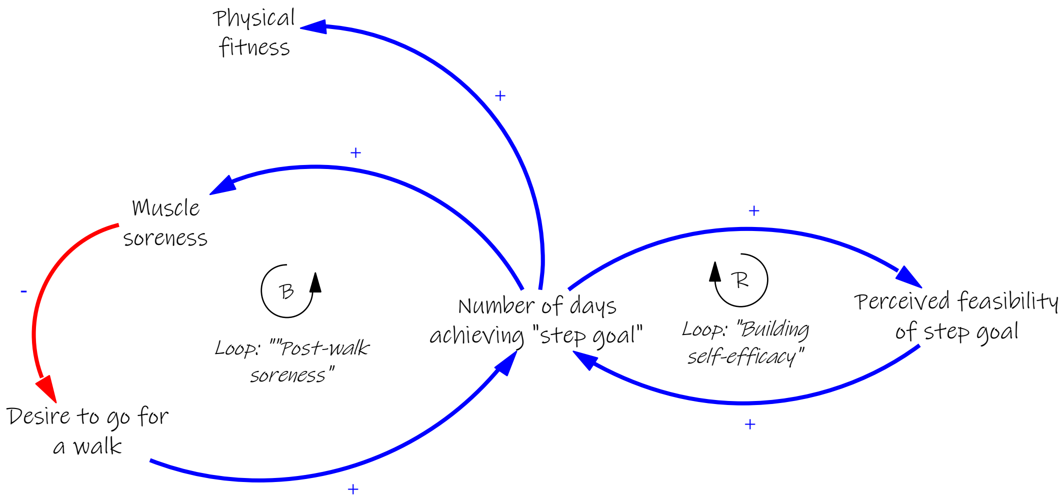

But, this effect might take a few weeks or even months to happen. We can mark delayed effects on the map using a tick mark as shown in this map.

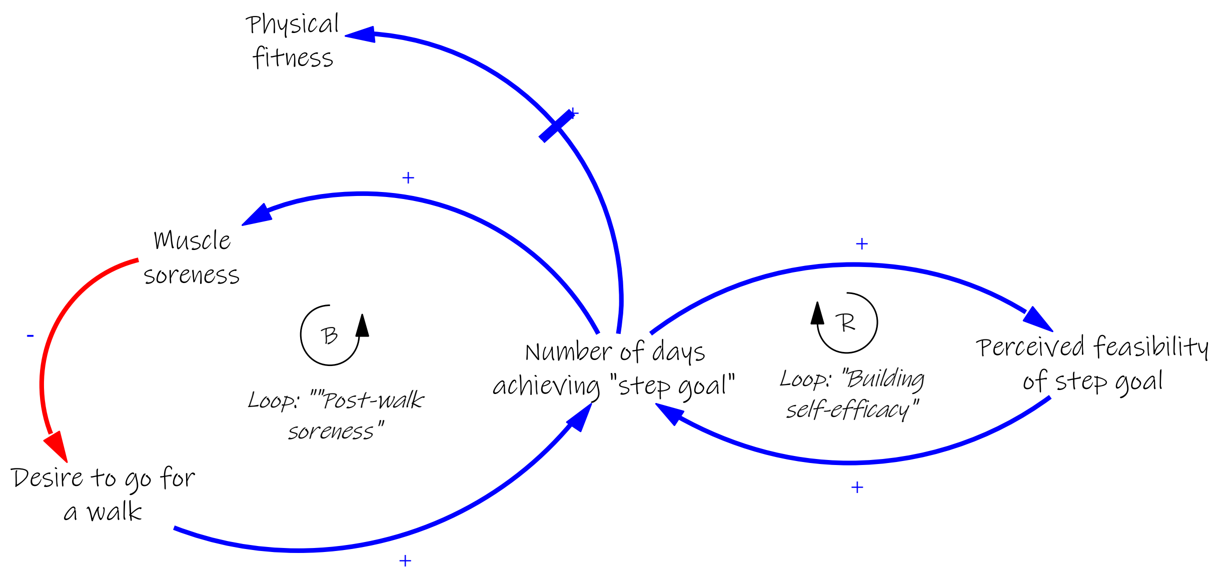


We can close this loop by drawing an arrow between physical fitness and soreness; this is a "-" arrow because as fitness increase, muscle soreness goes away.

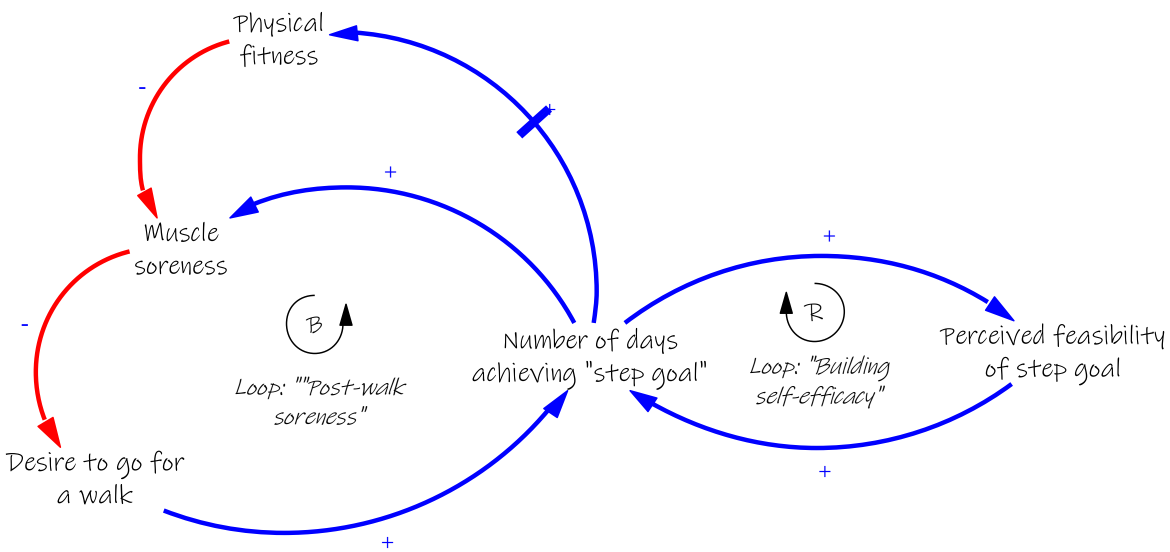


We can also draw an arrow from physical fitness to desire to go for a walk. This is a "+" arrow because walking may become more enjoyable as fitness increases. This creates another reinforcing loop, which we can label as "Getting in Shape."

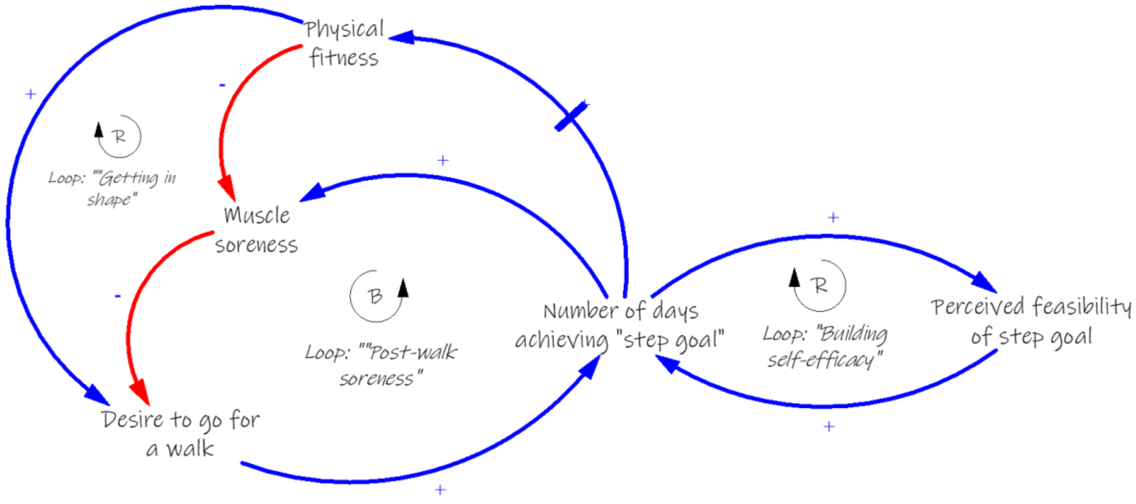

When we add the rest of the trends that we know were important to Betty and Jan, including knee pain, and other factors like walking buddies or cold weather, we can start to understand why this New Year's Resolution was actually quite complicated.

Here is a full system map:

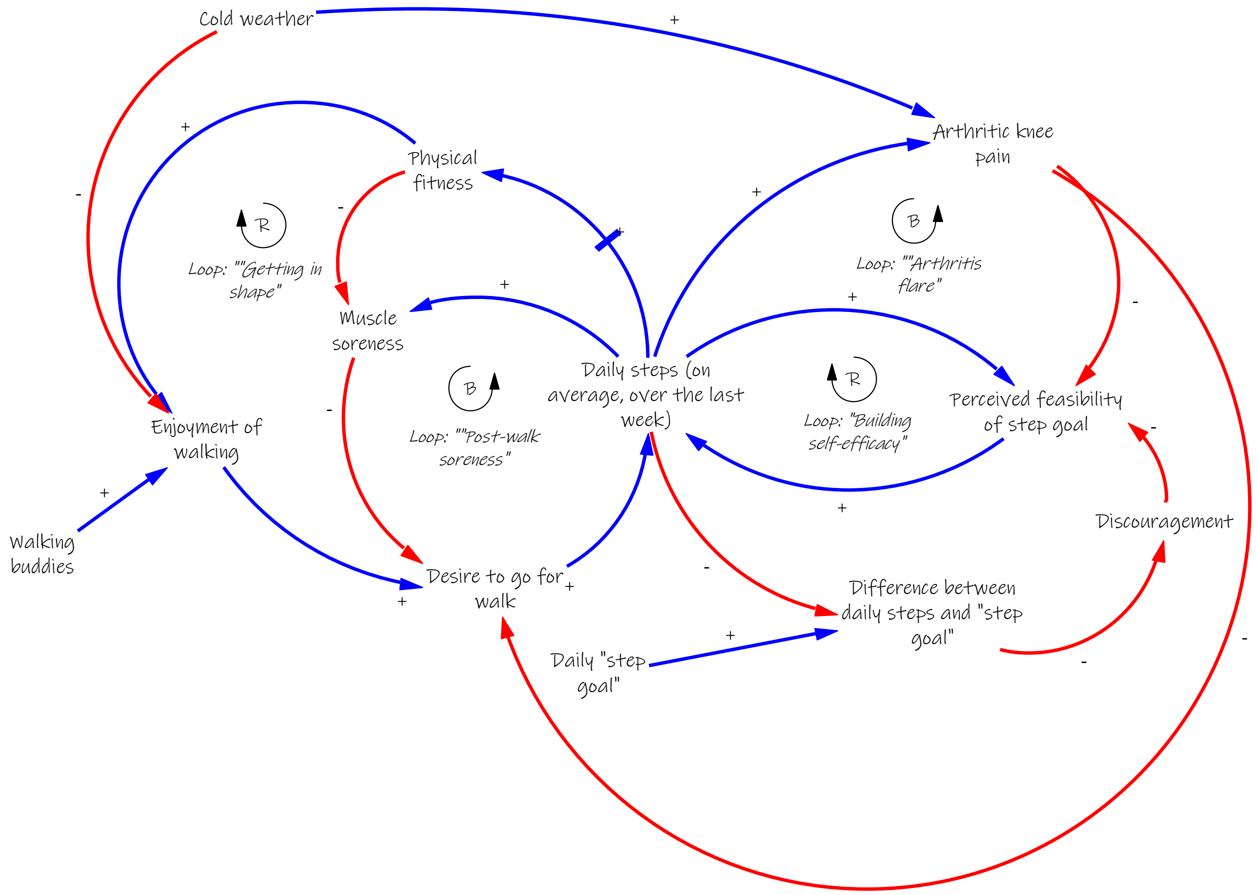

*Improving System Maps with New Information*

This map could also be expanded to include aspects of their environment or other pieces of context that different people may bring to this goal. For example, if Betty and Jan lived in very different places (in a city versus in the countryside, for example) that impacted how much they walked at baseline, we could include their this on the map.

We can also revise the maps as we learn more from people about their experiences with the same events or outcomes. Consider, for example, we met another person, Joe, who also wanted to walk 10,000 steps.

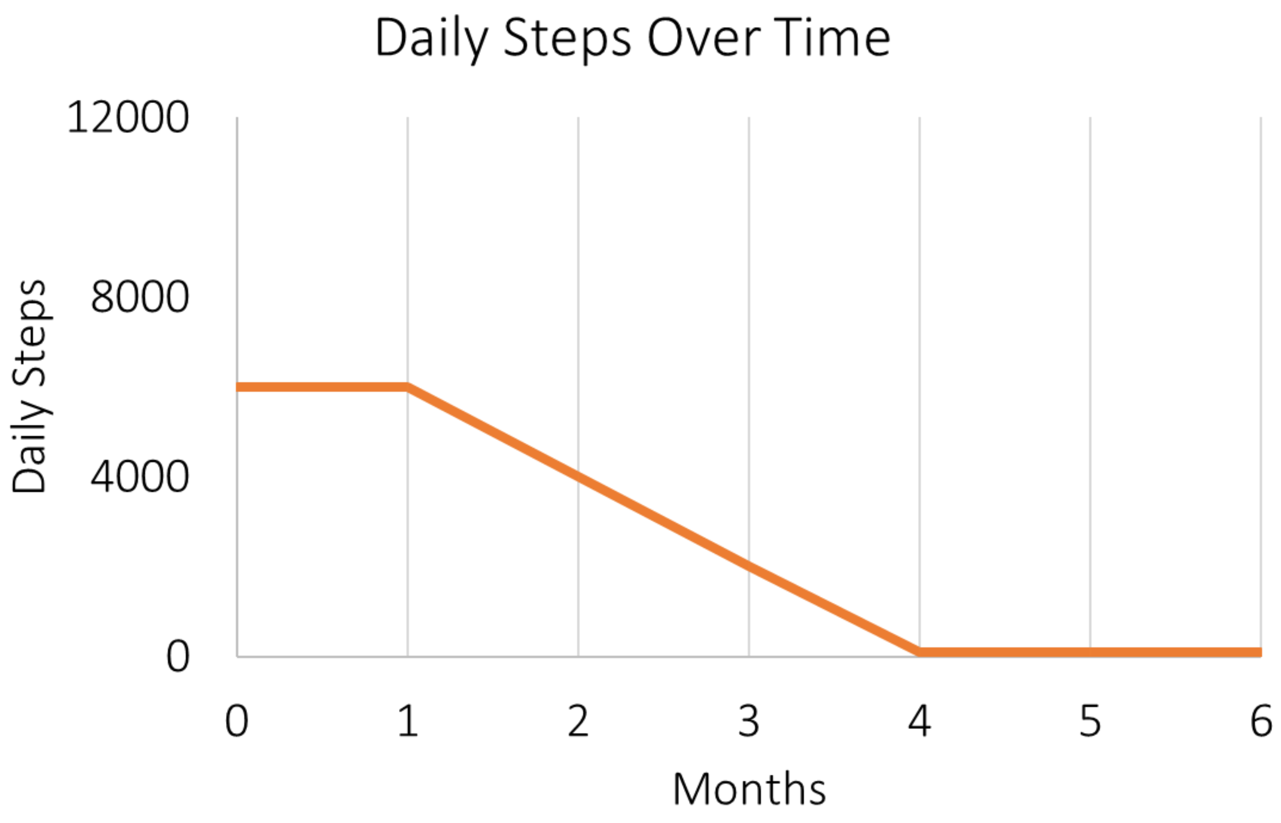

Here is his story:

Joe also set a resolution to walk 10,000 steps per day. However, this goal was too high for him. He became so discouraged early on that he did not walk at all.

We can add his experience into the map by drawing the relationship between the step goal, the difference between that goal and reality, and how it may impact on emotions (discouragement) and how feasible the goal may feel.

The revised map is below:

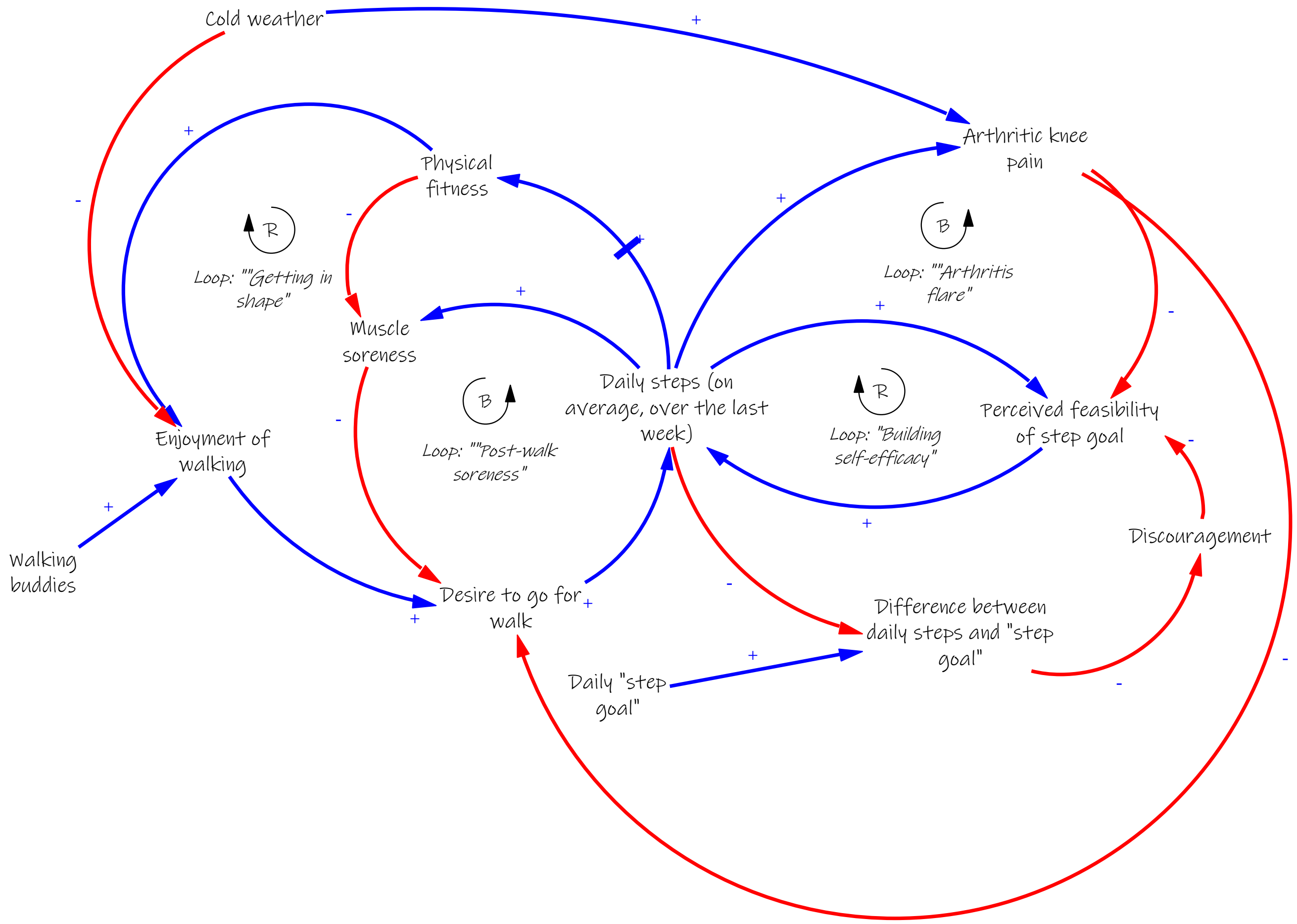

#### *3. How we can use systems thinking and system maps to better support patients achieving health and wellness*

We can use systems thinking, and specifically our new system map, to really see the full ‘iceberg’ that represents a person’s experience with their health, including the actions, behaviors, or habits that are often important for staying healthy but difficult to do.

As researchers, the system maps help us to understand how, and importantly, why different people may have different experiences. We can use these maps to explore and model different pieces and parts of the system.

As care providers, the system maps also show us where we can intervene to help.

For Betty, for example, we can use this map to see that we could better support her by identifying walking buddies sooner or helping her to acquire cold weather gear to stay warm. For Jan, we can see that we need to provide her with exercises and pain relief to avoid knee pain. Finally, for Joe, we can see that we need to adjust his starting step goal so it is more attainable and he does not get discouraged and quit.

The most important part for drawing system maps that are realistic and helpful for improving the way we offer medical care and support patients is to ensure that patients, providers, and caregivers can share their experiences, including all of the related factors, their patterns, and how goals and values affect what happens. It is only possible to dive below the waterline with all of these perspectives. It is also important that maps are revised so that they reflect new experiences and possible solutions for the complexity of staying healthy.

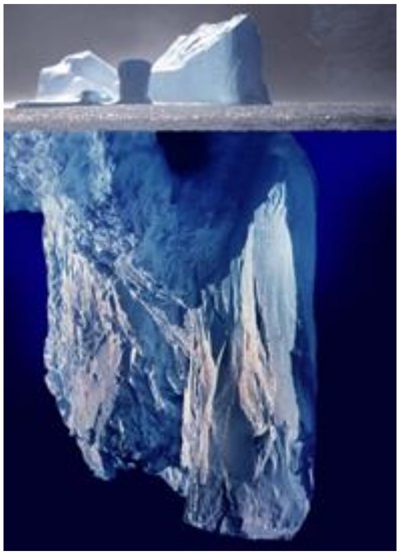

*The iceberg image is work by Uwe Kils http://www.ecoscope.com/iceberg/. Creative Commons CC BY-SA 3.0*

*Questions?*

*Please*

*Thank you for participating in the CGM Older Adult Stakeholder Mapping Workshop!*

**References:**

The 'Iceberg Model' for Systems Thinking adapted from: Kim, D. H. (1999). Introduction to systems thinking (Vol. 16): Pegasus Communications Waltham, MA. It has been used widely since then to teach systems thinking at an introductory level.

Parts of this guide were modeled after:

Frerichs L, Lich KH, Funchess M, Burrell M, Cerulli C, Bedell P, White AM. Applying critical race theory to group model building methods to address community violence. Progress in community health partnerships: research, education, and action. 2016;10(3):443.

The iceberg image is work by Uwe Kils http://www.ecoscope.com/iceberg/. Creative Commons CC BY-SA 3.0
The photo is from https://unsplash.com/s/photos/iceberg.
