## Supplementary material for "Use of Systems Thinking and Group Model Building Methods to Understand Patterns of Continuous Glucose Monitoring Use Among Older Adults with Type 1 Diabetes": A sample workshop packet from the latter five workshops can be found in Appendix B: Group Model Building Workshop Packet.

Appendix B: Sample Group Model Building Workshop Packet

### CGM Older Adult Stakeholder Mapping Workshop

#### Workshop Packet

Group Number _____ Participant Number _____

#### Activity #1: Warmup Exercise

Write down what you see as 3 most important things contributing to a positive experience for older adults when starting to use a CGM device to monitor blood sugar levels.

1.

2.

3.

Write down three things about using CGM that you think may be the most challenging parts for older adults.

1.

2.

3.

Group Number _____ Participant Number _____

#### Activity #2: Drawing CGM Use Patterns and Related Trends

##### What Kinds of Trends?

- There are no right or wrong answers
- Usually nouns (people, places, things, ideas, etc)
- Something that may increase or decrease over time
- Doesn't need to have a formal scale
- Can be consequences of causes of the "problem"

##### Ideas for Trends:

Blood sugar outcomes:

- Episodes of low blood sugar
- Episodes of severe low blood sugar
- Episodes of high blood sugar
- Time spent with high blood sugar
- Hypoglycemia unawareness
- HbA1c

Attitudes around blood sugar:

- Fear of low blood sugar
- Comfort with high blood sugar
- Self-confidence

Well-being outcomes:

- Satisfaction with glucose monitoring
- Diabetes distress
- Quality of life
- Anxiety
- Stress

Physical function or health:

- Fatigue
- Quality of sleep

CGM-specific experiences:

- Sensor, transmitter, receiver problems
- Painful to wear
- Inaccurate CGM readings or distrust in CGM readings
- Cost of supplies
- Challenges learning how to use new technology
- Frustration with learning how to use new technology
- Time spent learning new technology
- Ability to understand blood sugar readings and trends from CGM
- Value or usefulness of CGM
- Increased blood sugar readings to share
- Alarms and alerts

Group Number _____ Participant Number _____

**2A:** Which of these graphs most closely represents your own experience with CGM over the first six months?

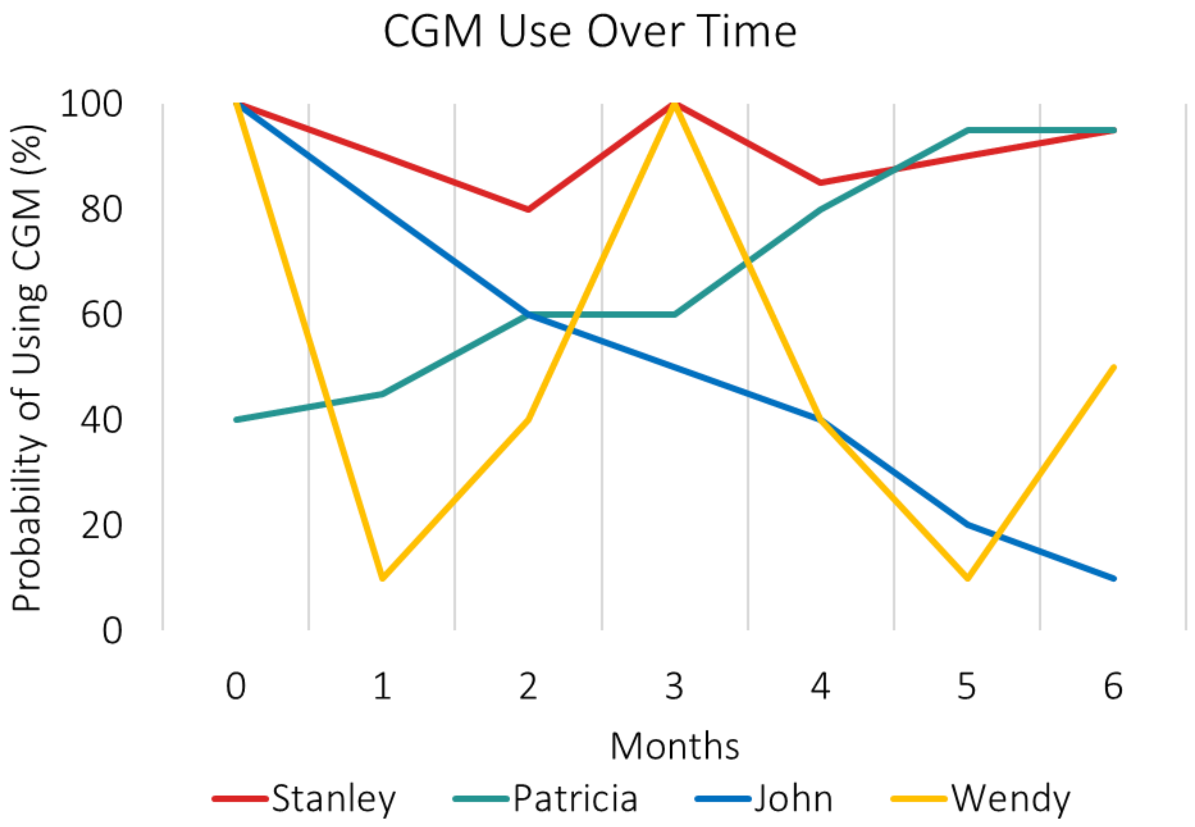


Your answer: ____________________________________

Now, use the next two pages to draw your own CGM use pattern over the first six months. (You should draw the same graph twice, we will need two copies).

Group Number _____ Participant Number _____

### My CGM Use (Graph 1)

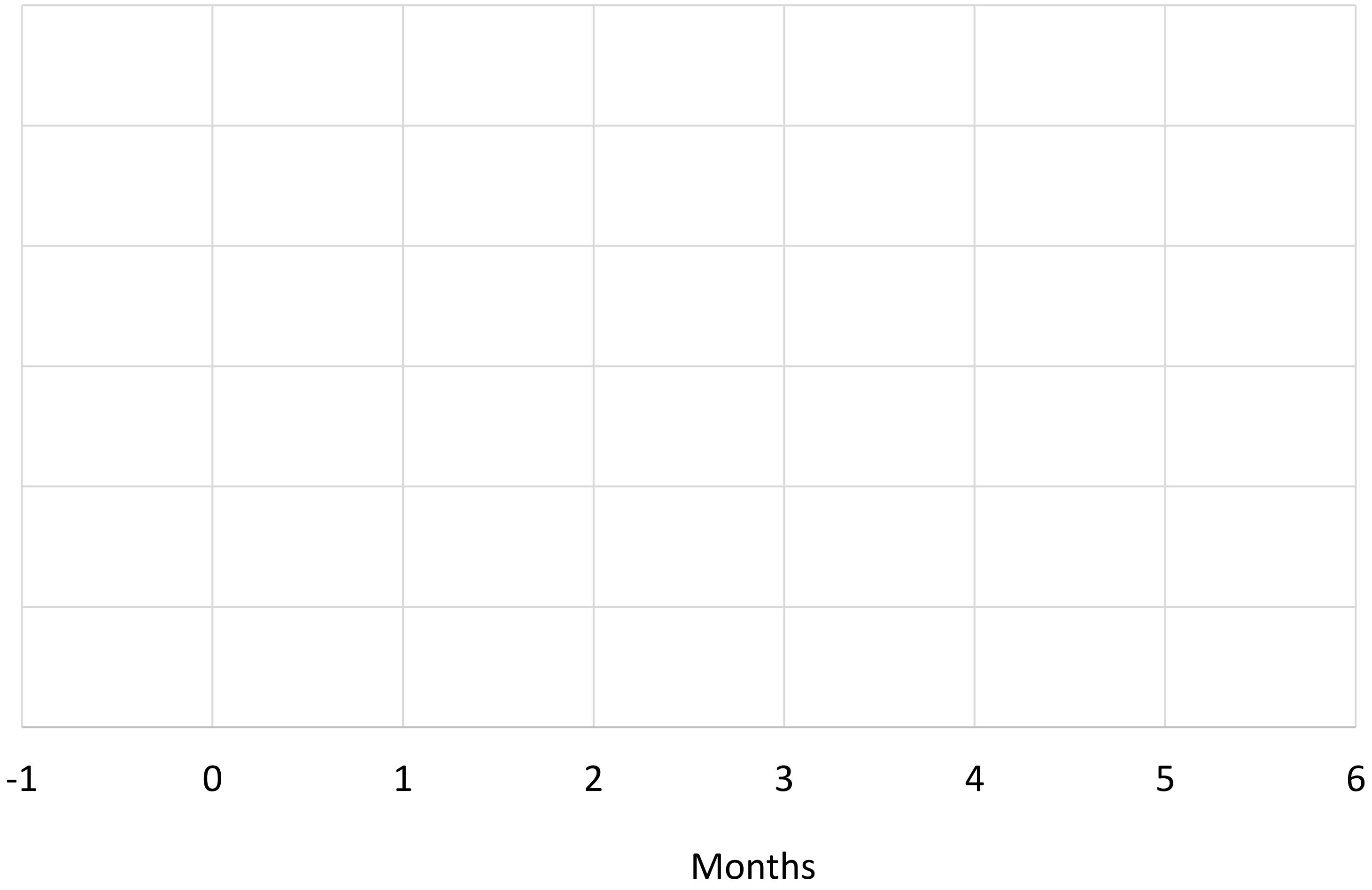


**2B:** What are three emotions that you felt in the first six months?

Add your emotions to the graph legend.

☐ Emotion #1: ___________________________

☐ Emotion #2: ___________________________

☐ Emotion #3: ___________________________

Then, draw a graph of those emotions over the first six months of using CGM.

Finally, mark and label three important events in the timeline that shaped your story.

Group Number _____ Participant Number _____

### My CGM Use (Graph 2)

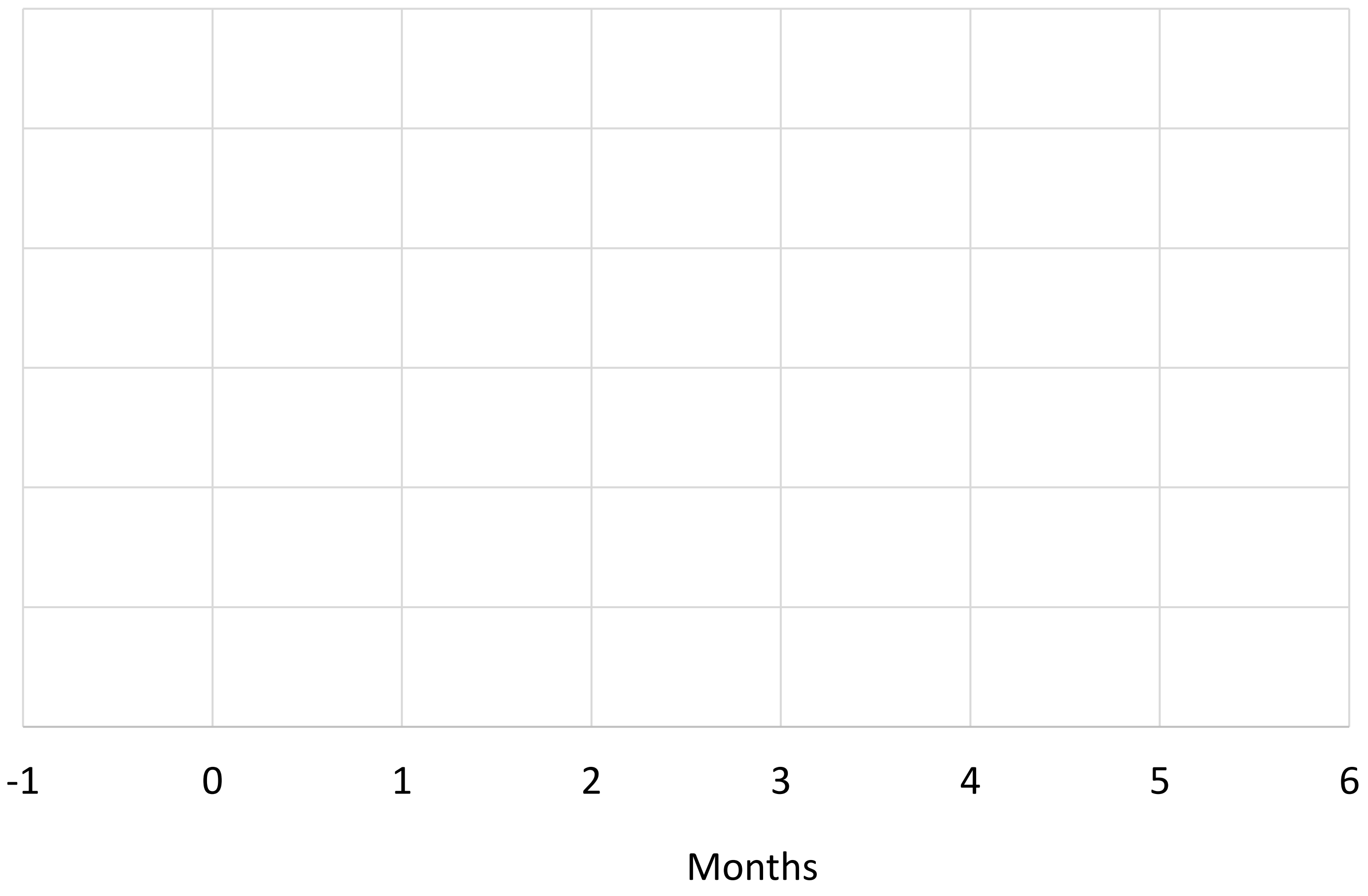


**2C:** We know different people find different benefits or values from using CGM. What are the three things that YOU value most about CGM? Use your second graph.

Add those benefits to the graph legend.

☐ Benefit #1: __________________________

☐ Benefit #2: __________________________

☐ Benefit #3: __________________________

Then, draw a graph of how those benefits looked over the first six months.

Finally, mark and label three important events in the timeline that shaped your story.

Group Number _____ Participant Number _____

Optional (and we will discuss): Since the first six months when you think about what has really shaped the experience, what are three things that you think are shaping your experience?

_____________________________________

_____________________________________

_____________________________________

Group Number _____ Participant Number _____

#### Final Reflections

1. During the workshop today, how comfortable did you feel to share all of your experiences and thoughts surrounding CGM use for older adults?

| Not comfortable at all | | Somewhat comfortable | | Very comfortable |
|---|---|---|---|---|
| 1 | 2 | 3 | 4 | 5 |

*Please explain why you felt this way:*

______________________________________________________________________________

______________________________________________________________________________

______________________________________________________________________________

2. Is there anything that you did not feel comfortable sharing with the group that you would like to share privately with the research team?

☐ No

☐ Yes

*If yes, please feel free to share here:*

______________________________________________________________________________

______________________________________________________________________________

______________________________________________________________________________

______________________________________________________________________________

______________________________________________________________________________

Group Number _____ Participant Number _____

3. What did you like about the workshop?

______________________________________________________________________
______________________________________________________________________
______________________________________________________________________
______________________________________________________________________
______________________________________________________________________

4. What did you not like about the workshop? Please provide suggestions for ways to improve the workshop.

______________________________________________________________________
______________________________________________________________________
______________________________________________________________________
______________________________________________________________________
______________________________________________________________________
______________________________________________________________________

Are you interested in seeing the final system maps that the research team creates based on the full series of workshops? You would be invited to give your feedback on the maps to the research team.

☐ Yes, please email me: ______________________________

☐ No

**Thank you!**
